# Applying a multiplex electrochemiluminescence NS1 assay to detect and differentiate Zika and dengue virus exposures in long-term community cohorts in Brazil and Thailand

**DOI:** 10.64898/2026.09.04.26362297

**Authors:** Nivison Nery, Marco Hamins-Puertolas, Lauren Bahr, Jaqueline S. Cruz, Joseph Lu, M. Catherine Muenker, Juan P. Aguilar Ticona, Ruchira Khosavanna, Mariam O. Fofana, Daiana de Oliveira, Darunee Buddhari, Taweewun Hunsawong, Aaron Farmer, Kathryn A. McGuckin, John S. Brooks, Sopon Iamsirithaworn, Matthew H. Collins, Guilherme S. Ribeiro, BUZZ Study Team, Stephen J. Thomas, Federico Costa, Mitermayer G. Reis, Derek A.T. Cummings, Kathryn Anderson, Albert I Ko, Adam Waickman

## Abstract

**Background:** Distinguishing Zika virus (ZIKV) from Dengue virus (DENV) infection remains a major diagnostic challenge in co-endemic regions due to antigenic cross-reactivity. We evaluated a multiplex electrochemiluminescence Meso Scale Discovery System (MSD) NS1 assay to define ZIKV infection with high specificity across diverse immune backgrounds.

**Methods:** We analyzed longitudinal samples from PCR-confirmed ZIKV and DENV infections across multiple cohorts, assessed correlations between MSD NS1 signal intensities and PRNT_50_ titres, and performed ROC analyses to derive seroconversion thresholds. We then applied these criteria to identify ZIKV seroconversions in Brazil (2015-2024) and Thailand (2015-2025)

**Findings:** ZIKV infections produced sustained NS1 responses, whereas primary DENV infections showed minimal ZIKV cross-reactivity and secondary DENV infections exhibited only transient, low-level increases. MSD NS1 signal intensities correlated with PRNT_50_ titres for ZIKV and all four DENV serotypes (ρ = 0.54–0.84). ROC analyses identified a fold-change ≥ 10.96 as the optimal ZIKV seroconversion cutoff, with an AUC of 100% among DENV-naïve and 90.7% among DENV-exposed participants. Adding a second criterion — ZIKV/DENV fold-change ratio > 1 — further increased specificity. Application of this dual-threshold framework revealed high ZIKV incidence during the 2015–2016 epidemic in Brazil and very low incidence in the post-epidemic period. In the Thailand cohort, synchronous seroconversion among household members suggested potential intra-household transmission.

**Interpretation:** The MSD NS1 multiplex assay provides a scalable, quantitative, and highly specific approach for identifying ZIKV infection and estimating seroincidence in co-endemic settings. The dual-threshold framework (fold-change ≥ 10.96 and ZIKV/DENV ratio > 1) enhances diagnostic precision, offering a robust tool for surveillance and transmission studies in flavivirus-endemic regions.

**Funding:** Wellcome Trust Fund; The National Council for Scientific and Technological Development (CNPq; grant: 440891/2016–7, 311365/2021–3, and 303307/2026-9 to G.S.R.); the Bahia Foundation for Research Support (grant PET0022/2016 to G.S.R.); the Coordination for the Improvement of Higher Education Personnel, Brazilian Ministry of Education (grant 88881.130749/2016–01 to G.S.R.)

**RESEARCH IN CONTEXT:** *Evidence before this study:* We searched PubMed, Scopus, and Web of Science for articles published in English up to May 1, 2026, using combinations of the terms “Zika virus”, “dengue virus”, “NS1”, “serology”, “cross-reactivity”, “electrochemiluminescence”, “multiplex assay”, “seroincidence”, and “PRNT”. Previous studies demonstrated that serological discrimination between Zika virus (ZIKV) and dengue virus (DENV) infections is challenging because of extensive flavivirus cross-reactivity, particularly in DENV-endemic settings. Conventional ELISA-based assays frequently lack specificity, especially among individuals with prior flavivirus exposure. Plaque reduction neutralization testing (PRNT) remains the reference standard for differentiating flavivirus infections, but its low throughput, high cost, and technical complexity limit its application in large epidemiological studies. Recent studies have highlighted NS1-based assays as promising alternatives because NS1 antibodies appear more virus-specific than envelope-directed responses. Multiplex electrochemiluminescence platforms, including Meso Scale Discovery (MSD), have shown high analytical sensitivity and scalability for infectious disease serology, but evidence supporting their use for distinguishing ZIKV from DENV infections across distinct immune backgrounds and longitudinal cohort settings remains limited. Few studies have evaluated quantitative seroconversion thresholds for identifying ZIKV infection in populations with prior DENV exposure or explored their application for long-term seroincidence estimation and transmission inference.

*Added value of this study:* This study validates a multiplex electrochemiluminescence NS1 assay for differentiating ZIKV and DENV infections using multiple well-characterized cohorts from Brazil, Thailand, and the United States, including PCR-confirmed ZIKV and DENV infections, longitudinal community cohorts, and a controlled human dengue infection model. We identified a standardized serological threshold based on ZIKV NS1 fold-change (≥10·96) and demonstrated that combining this threshold with a second criterion—the ZIKV-to-DENV fold-change ratio > 1, substantially improves specificity in highly DENV-endemic populations. We additionally showed strong correlations between MSD signal intensities and PRNT_50_ neutralization titres for ZIKV and all four DENV serotypes, supporting the use of the assay as a scalable surrogate for neutralization-based approaches. Application of this dual-threshold framework enabled estimation of ZIKV seroincidence over nearly a decade of follow-up in Brazil and identified patterns suggestive of intra-household transmission in Thailand. Together, these findings provide a practical high-throughput strategy for distinguishing flavivirus infections in epidemiological and surveillance settings.

*Implications of all the available evidence:* Accurate discrimination between ZIKV and DENV infections remains a major barrier for flavivirus surveillance, seroepidemiological studies, and vaccine evaluation in co-endemic regions. Our findings suggest that multiplex NS1 electrochemiluminescence assays combined with quantitative dual-threshold criteria can provide a scalable and standardized alternative to PRNT for identifying ZIKV infections in populations with heterogeneous flavivirus exposure histories. This framework could strengthen surveillance systems by improving estimates of ZIKV seroprevalence and seroincidence, supporting detection of silent or low-level transmission, and enabling more reliable interpretation of longitudinal immune responses in endemic settings. The approach may also facilitate vaccine studies and outbreak investigations in regions where multiple flaviviruses co-circulate.

**Key messages:**

1. **Strong and long-lasting ZIKV NS1 responses with minimal DENV cross-reactivity** The Meso Scale Discovery System (MSD) multiplex NS1 assay measured robust and durable ZIKV-specific antibody responses following PCR-confirmed infection, while primary and secondary DENV infections produced only minimal or transient ZIKV NS1 reactivity, demonstrating high antigenic specificity.
2. **MSD NS1 signals correlate strongly with PRNT_50_ neutralization titres** Across multiple cohorts, MSD signal intensities showed strong concordance with PRNT_50_ titres for both ZIKV and DENV serotypes, validating the MSD assay as a reliable, scalable surrogate for functional neutralization assays.
3. **A dual-threshold framework improves diagnostic accuracy** ROC analyses identified a fold-change ≥ 10.96 as a sensitive and specific marker of ZIKV seroconversion, further refined by a ZIKV/DENV fold-change ratio > 1 to enhance specificity and reduce misclassification in DENV-exposed populations.
4. **Accurate detection of ZIKV seroincidence and transmission patterns** Application of the dual-threshold criteria revealed high ZIKV incidence during the 2015–2016 epidemic in Brazil and very low transmission in subsequent years. In a Thai household, spatiotemporal clustering of seroconversion among three members is suggestive of intra-household transmission, demonstrating the framework’s utility for detecting potential transmission clusters in seroepidemiological surveillance.

## Introduction

Zika virus (ZIKV) and dengue virus (DENV) are closely related flaviviruses that co-circulate in many tropical and subtropical regions, particularly in Latin America and Southeast Asia. Both viruses share the same primary mosquito vectors (*Aedes aegypti* and *Aedes albopictus*) and exhibit overlapping clinical manifestations—such as fever, rash, arthralgia, and conjunctivitis—which complicates syndromic diagnosis in endemic settings^1^. Although the explosive ZIKV epidemic in the Americas subsided after 2016, accumulating evidence indicates that transmission continues at lower and often undetected levels in multiple regions^2–5^, making accurate laboratory differentiation essential given the distinct clinical outcomes associated with each virus, most notably the link between ZIKV infection and congenital Zika syndrome as well as neurological complications^6–9^.

A major challenge in understanding the true burden and transmission dynamics of ZIKV is the serological discrimination between ZIKV and DENV infections. Due to their close antigenic relatedness, conventional assays such as ELISA frequently yield cross-reactive results, obscuring the true burden of infection and complicating estimates of seroprevalence and seroincidence in settings where these viruses co-circulate^10–13^. This challenge is particularly pronounced in populations with prior flavivirus exposure, where sequential infections, flavivirus vaccination history, and long-lived cross-reactive antibodies complicate interpretation of immune responses^14–17^. Although the plaque reduction neutralization test (PRNT) remains the gold standard for distinguishing flavivirus infections, its labor-intensive protocol, low throughput, and high cost render it impractical for large-scale cohort studies^13^. In addition, PRNT interpretation remains challenging in populations with prior flavivirus exposure or vaccination as cross-neutralizing antibodies can reduce virus-specific discrimination^18^. To address these limitations, new serological tools are urgently needed.

Standardized methodologies for serosurveys, stratified by age cohorts and based on assessments of serological assays, would serve to elucidate patterns of Zika virus transmission worldwide ^19–21^. Multiplex electrochemiluminescence platforms, such as the Meso Scale Discovery (MSD) system, offer a high-throughput and scalable alternative for serological surveillance^22^. This platform enables simultaneous detection of antibodies against multiple viral antigens with high sensitivity and specificity along with a large dynamic range^23^. MSD-based assays have been successfully applied to quantify antibody responses to SARS-CoV-2 and other pathogens and have shown strong correlation with neutralization assays^22^. These characteristics make the MSD platform a promising approach for distinguishing flavivirus infections and detecting otherwise unrecognized ZIKV transmission in co-endemic settings.

In this study, we evaluated the performance of a multiplex MSD NS1 assay to differentiate ZIKV from DENV infections in two well-characterized community cohorts: (i) the Pau da Lima cohort in Salvador, Brazil which provided a unique opportunity to assess antibody responses before, during, and up to eight years after the 2015 ZIKV epidemic; and (ii) the Kamphaeng Phet family cohort in Thailand, a prospective multigenerational family-based cohort under active dengue surveillance, which enabled assessment of assay performance in differentiating DENV infections from ZIKV infection in a hyperendemic Southeast Asian setting. Together, these analyses support the use of the MSD multiplex assay for seroprevalence and seroincidence studies of ZIKV and DENV in populations with distinct backgrounds of flavivirus exposure.

## Methods

We leveraged samples from multiple sources to collectively address distinct aspects of assay validation: (i) community-based cohorts with longitudinal sampling to assess seroincidence and transmission patterns; (ii) well-characterized patient cohorts with PCR-confirmed infections to validate assay performance against a gold standard; and (iii) controlled infection models to evaluate specificity in the absence of confounding exposures. We analyzed serum and plasma samples from longitudinal community-based cohorts in Brazil and Thailand, which provided natural exposure settings with available paired (pre- and post-infection) samples with inferred and confirmed exposure to ZIKV and DENV, respectively. Both settings are DENV endemic, where distinguishing ZIKV from DENV is particularly challenging due to the widespread DENV immunity and flavivirus vaccination coverage (Yellow Fever and Japanese Encephalitis in Brazil and Thailand respectively)^24–26^. In addition, we included samples from three well-characterized patient cohorts–clinic based Brazilian patients with confirmed ZIKV infections, recent travelers with confirmed ZIKV infection in the United States (U.S.), and participants from a controlled human dengue virus infection model (DHIM). Together, these cohorts enabled a comprehensive assessment of assay specificity, cross-reactivity, and immune response dynamics across diverse populations.

### Longitudinal community cohorts

#### Brazil (Pau da Lima cohort)

We analyzed samples from a community-based prospective cohort established in Pau da Lima, a densely populated urban neighborhood in Salvador, Brazil. The cohort was established in 2014 for arboviral surveillance and has been followed through 2024. During the Zika epidemic of 2015, approximately 70% of the population was estimated to have been exposed to ZIKV using serological approaches^27^. For this study, we selected 268 participants with available serum samples collected before (2014), during (2015–2016), and after the epidemic, with up to 8 years of longitudinal follow-up. Participants were classified according to serological evidence of prior DENV exposure based on pre-epidemic samples.

#### Thailand (Kamphaeng Phet cohort)

The second cohort for this analysis was derived from a long-standing community-based dengue study established in 2015 in Kamphaeng Phet, Thailand^28^. This cohort allows for the evaluation of assay performance in another setting with high DENV endemicity. For the present analysis, we included paired and longitudinal serum samples from 85 participants (median age of 12, range of 1-30) where either they or a household member had a PCR-confirmed DENV infection during the interval of interest, together with available pre-infection baseline samples. We leveraged these PCR-confirmed DENV infections to characterize the antibody responses to MSD NS1 assays in non-ZIKV flavivirus infections.

### Patient cohorts

#### Brazil Zika cases

We included serum samples from 19 participants enrolled in an acute febrile illness surveillance study designed to detect arbovirus infections during the 2015–2016 Zika virus (ZIKV) outbreak in Salvador, Brazil^29^. These individuals presented with clinical manifestations consistent with Zika infection and were confirmed to be ZIKV positive by PCR. Samples were collected during the acute and convalescent phases of illness^5^. This cohort was used to characterize the antibody responses and evaluate the performance of MSD NS1 assays in confirmed ZIKV infections within a Brazilian population context.

#### US Zika cases

The cohort consisted of U.S. participants who presented with symptoms, had recent travel history (or reported sexual contact with a recent traveler with symptomatic illness), and were clinically evaluated at Emory University^30^. Participants had documented ZIKV exposure confirmed by PCR and/or serology, with participants likely to be flavivirus- naïve prior to infection, enabling assessment of ZIKV-specific antibody responses in the absence of confounding DENV immunity. Acute and follow-up serum samples were analyzed to assess antibody kinetics and assay specificity in seven individuals with well-defined infection timing and travel-related exposure history.

#### Dengue human infection model (DHIM

Samples were obtained from 14 participants enrolled in a controlled human dengue virus infection model conducted in the U.S. In this study, flavivirus-naïve adult volunteers were inoculated with a well-characterized DENV-1, -3, or -4 strain and monitored prospectively for clinical symptoms and immune responses ^31–34^. This cohort provided a controlled setting for evaluating assay specificity where any ZIKV NS1 reactivity following DENV-1 infection would represent true cross-reactivity rather than pre-existing immunity. We used serum samples from the pre-infection, acute, and convalescent phases to evaluate the specificity of MSD NS1 responses to DENV and potential cross-reactivity with ZIKV antigens.

### Ethics statement

The surveillance study for arboviral infections among participants with acute febrile illness was approved by the Institutional Review Board of the Oswaldo Cruz Foundation (Salvador, Brazil), and written informed consent was obtained from all participants.

The U.S. Traveler study was reviewed and approved by the institutional review board at Emory University.

The DHIM and associated analysis were approved by the State University of New York Upstate Medical University (SUNY-UMU) and the Department of Defense’s Human Research Protection Office.

The Pau da Lima cohort protocol was approved by the Institutional Review Board of the Oswaldo Cruz Foundation, Salvador, Brazil, and Yale University.

The Kamphaeng Phet cohort protocol was approved by the Institutional Review Board of the Thai Ministry of Public Health and collaborating institutions.

Written informed consent (and assent for children) was obtained from all participants or their guardians.

### Multiplex electrochemiluminescence assay (MSD SU NS1)

Serum samples were tested using a multiplex electrochemiluminescence assay (Meso Scale Discovery, Rockville, MD) based on NS1 antigens. The custom MSD NS1 panel included recombinant NS1 proteins from six flaviviruses: ZIKV (Suriname), DENV1–4 (Nauru/Western Pacific/1974, Thailand/16681/84, Sri Lanka D3/H/IMTSSA-SRI/2000/1266, Dominica/814669/1981), and Japanese encephalitis virus (JEV, SA-14) with titres reported as signal units (SU). Additional information regarding the assay protocol and reproducibility can be found in the Supplemental.

### Comparator assays

#### Plaque Reduction Neutralization Test (PRNT)

PRNT was performed on a subset of samples at reference laboratories using standardized protocols and was tested for DENV1-4 (BR-PE/97-42735, BR-PE/95-3808, BR-PE/95-3808, BR-PE/12-008) and ZIKV (BR-PE243/2015) in Brazil while it was tested for DENV1-4 (Thailand/16007/1964, Thailand/16681/1984, Philippines/16562/1964, Thailand/C0036/2006) and JEV (SA-14-14 vaccine strain) in Thailand^27,35^. A subject was considered seropositive when their serum was able to reduce the number of viral plaques by at least 50% (PRNT_50_) compared with virus controls, at a dilution of >1:100 for ZIKV and 1:10 or higher, for any of the four DENV serotypes.

#### IgG3 antibody responses

A subset of Brazilian samples was tested for ZIKV-specific IgG3 response, which has been proposed as a biomarker of recent ZIKV infection^27^. ELISA was performed using recombinant ZIKV antigens, and IgG3 levels were quantified by optical density (OD) values.

### Statistical analysis

Spearman’s correlation coefficients were used to assess concordance between MSD SU values, PRNT titres, and IgG3 levels. Antibody kinetics over time were modeled using mixed-effects linear regression with random intercepts for participants.

To define the serological threshold that best discriminates ZIKV infection from non-infection, we evaluated the performance of the ZIKV NS1 antigen response measured by the MSD multiplex electrochemiluminescence assay. Fold-change values were calculated for each participant as the ratio of the ZIKV NS1 signal measured during versus before the 2015 ZIKV epidemic (i.e., [ZIKV_during_/ZIKV_pre_], where ZIKV_during_ and ZIKV_pre_ are the ZIKV specific MSD antibody titres during and after the 2015 ZIKV epidemic). Analyses were restricted to participants with available paired samples and known infection status based on ZIKV IgG3 seroconversion, defined as the appearance of IgG3 antibodies against ZIKV NS1 in post-epidemic samples. Baseline serological data was not available for samples (U.S. travelers and Brazil Zika cases) with PCR- confirmed ZIKV infection making these an inadequate reference standard for this analysis.

Two complementary case definitions were applied to classify ZIKV infection events using DENV1-4 and ZIKV antibody responses:

1. Primary fold-change definition: participants were considered positive if the ZIKV NS1 fold change (post-/pre-epidemic) was ≥ 10.96 (or equivalently ≥10.96 fold change), corresponding to the threshold identified in the following ROC-based analyses of seroconversion.

2. Relative fold-change definition: participants were considered positive if their ZIKV NS1 fold-change ratio was greater than the maximum fold-change observed among DENV 1–4 NS1 response. This criteria hypothesizes that ZIKV responses to ZIKV infection exceed cross-reactive dengue responses within the same individual.

To identify the optimal discrimination threshold, we performed receiver operating characteristic (ROC) curve analyses using the pROC package in R (version 4.3.3). We constructed ROC curves by plotting sensitivity (true positive rate) against 1 – specificity (false positive rate) across all possible cut-off values of the ZIKV NS1 fold change. The area under the curve (AUC) was used to quantify the overall diagnostic performance of the MSD assay in differentiating ZIKV IgG3 seroconverters (true positives) from non-seroconverters (true negatives). We used ZIKV-specific IgG3 seroconversion as the reference standard because IgG3 responses to ZIKV NS1 have been shown to be highly specific and minimally cross-reactive with DENV^10^.

To assess threshold robustness, we performed a 10-fold cross-validation approach. The optimal fold-change threshold was determined using Youden’s J index, calculated as *(sensitivity + specificity – 1)*, which identifies the cut-off that maximizes both sensitivity and specificity. Separate ROC analyses were performed for DENV-exposed and DENV-naïve individuals to evaluate potential differences in assay performance associated with pre-existing flavivirus immunity. This approach verified that threshold selection was stable across different subsets of the data. The final threshold applied to the full dataset was determined using all available samples.

To visualize confirmed DENV infections and ZIKV seroconversions, we performed principal component analysis (PCA) using the `prcomp` function from the `stats` package in R. The PCA incorporated the fold change (ratio of post-infection to pre-infection MSD signal) for each DENV serotype, ZIKV, and JEV. We used fold changes rather than raw MSD signal values to ensure that separation in the PCA space was driven by relative increases in antibody response due to infection events, rather than by baseline immunological differences between the Thailand and Brazil populations.

## Results

### Dynamics of ZIKV- and DENV-specific antibody responses (Figure 1)

We assessed longitudinal antibody responses in individuals with virologically (US patient cohorts) and PCR-confirmed (Brazil) ZIKV infections as well as PCR-confirmed DENV infections (Thailand cohort and DHIM) to validate MSD assay performance against gold-standard confirmation methods (Figure 1, Table S1).

**Figure 1.**
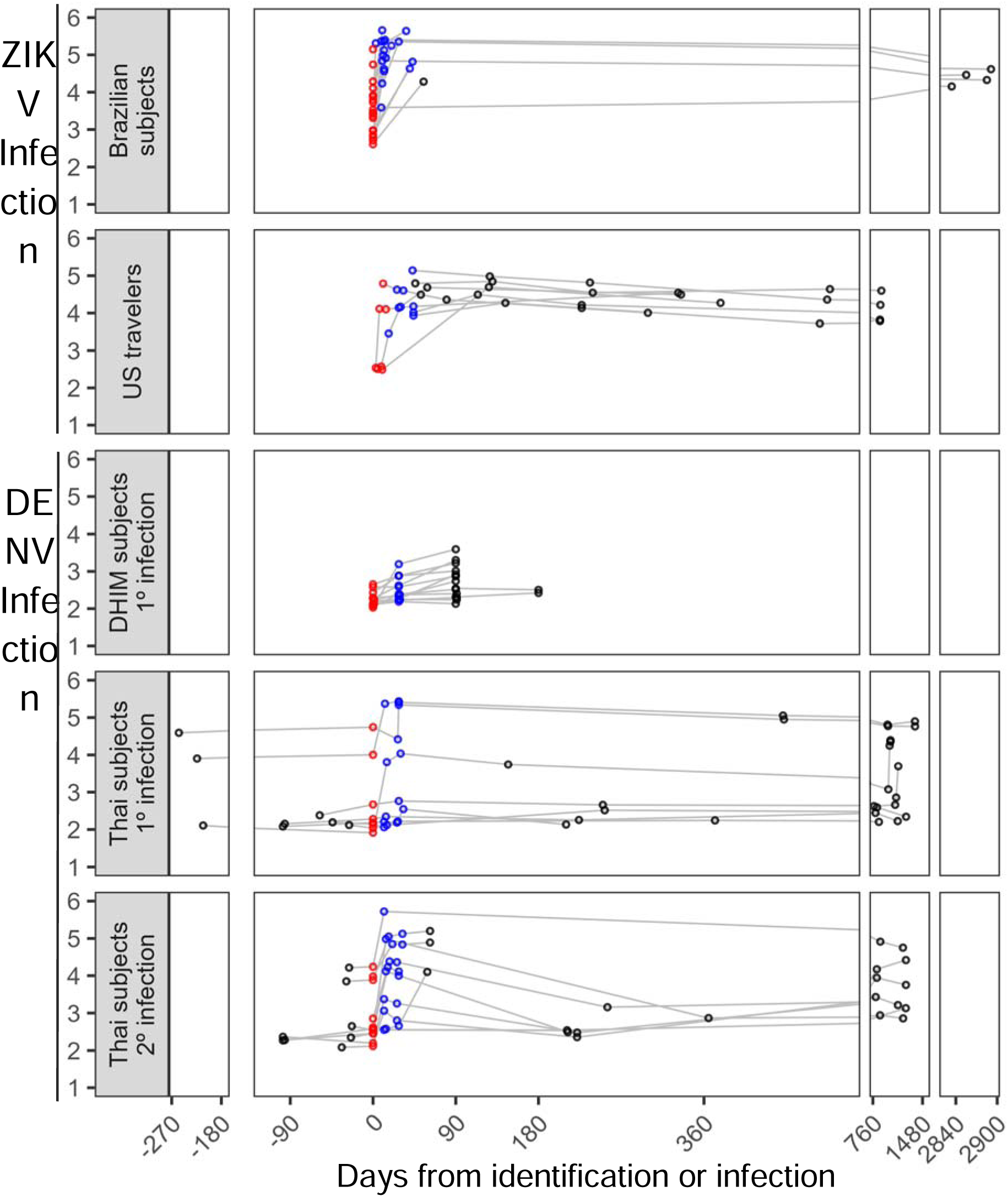
Dynamics of anti-ZIKV and anti-DENV NS1 antibodies after infection. Longitudinal antibody responses measured by MSD assay across different cohorts. ZIKV infections are from Brazilian subjects and U.S. travelers while DENV infections are from Dengue Human Infection Model participants with primary infection, and Thai subjects with primary and post-primary infections. Antibody levels (y-axis, log scale) are plotted over time since infection or case identification (x-axis, in days). Red symbols indicate acute-phase samples, blue symbols represent early convalescent samples, and black symbols correspond to later follow-up samples. The durable elevation of ZIKV NS1 responses after confirmed ZIKV infection and comparatively minimal elevations in DENV-infected participants supports the assay’s specificity in flavivirus-endemic settings.

Among ZIKV-infected individuals from Brazil and the United States, ZIKV NS1 responses rose sharply in the acute phase and remained elevated for several years post-infection. Signal intensities increased rapidly, with many participants exceeding 10⁵ SU within weeks of infection and showed sustained reactivity throughout follow-up (8 years), consistent with robust and long-lasting serological responses to ZIKV^36^.

In contrast, individuals with primary (defined as individuals either enrolled as newborns or seronegative upon enrollment and until infection event) DENV infection (Thailand, neonates) exhibited only minimal cross-reactivity to ZIKV NS1, with SU signals remaining near background levels. In contrast, those with post-primary DENV infections showed transient or low-level increases in ZIKV NS1 reactivity, but the majority of responses declined over time and remained well below those observed in confirmed ZIKV infections.

In the dengue human infection model, no appreciable increase in ZIKV NS1 signals were observed, further supporting the specificity of the MSD assay for distinguishing ZIKV from DENV infection.

Together, these results demonstrate that the MSD SU ZIKV NS1 assay yields strong and durable antibody responses following PCR-confirmed ZIKV infection, maintaining minimal cross-reactivity in primary DENV infections, while early heterogeneity among secondary DENV infections is exhibited during the early convalescent period (Figures S1-2).

### Correlation of MSD and PRNT_50_ (Figure 2)

We assessed correlations between MSD NS1 signal intensities and neutralization titres (PRNT_50_) across all cohorts (Figure 2). For ZIKV, MSD NS1 responses were strongly correlated with PRNT_50_ titres in the Brazil cohort (ρ = .83, p < 0.0001) and moderately correlated in the Thailand cohort (ρ = .54, p < 0.001). For DENV1–4, MSD NS1 responses also demonstrated positive correlations with PRNT_50_ values across cohorts. In Brazil, Spearman’s ρ ranged from .64 (DENV4) to .84 (DENV3), whereas in Thailand, correlations ranged from .68 (DENV2) to 0.79 (DENV3), all statistically significant (p < 0.0001).

**Figure 2.**
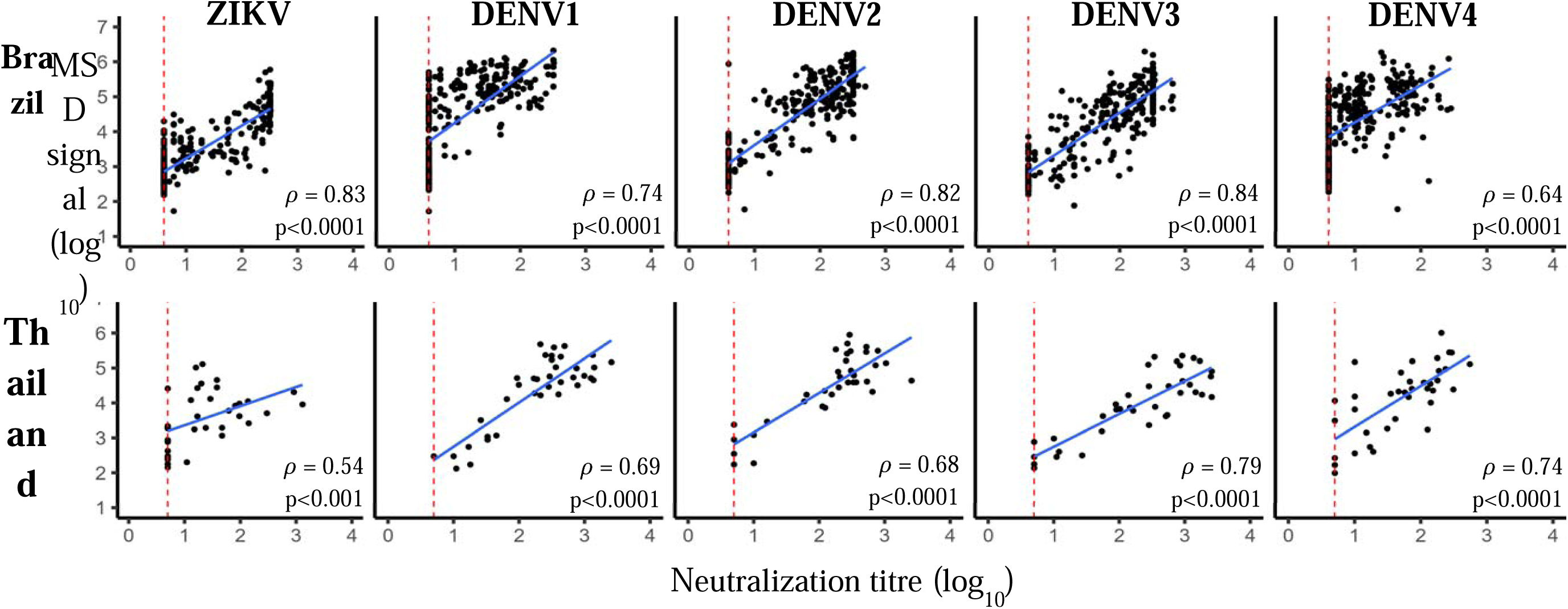
Correlation between MSD and PRNT results for ZIKV and DENV (serotypes 1–4) in Brazil and Thailand cohorts. MSD signal (log₁₀) values are plotted against PRNT titres (log₁₀) for each antigen. Blue lines represent regression fits, and ρ indicates Spearman’s correlation coefficient. Red dashed lines show the negative cut-off point for PRNT (Brazil: 4; Thailand: 5) and for IgG3 (0.586). Strong positive correlations were observed across ZIKV and DENV antigens, supporting the reliability of MSD for serological assessment.

Overall, these analyses demonstrate that the MSD multiplex NS1 assay provides a reliable and quantitative surrogate for PRNT_50_ measurements, accurately reflecting serotype-specific and virus-specific neutralizing antibody responses to both ZIKV and DENV in populations with diverse immune backgrounds.

### Application of MSD assay to seroincidence studies and definition of ZIKV infection threshold (Figure 3)

The MSD multiplex NS1 assay demonstrated high accuracy in identifying ZIKV infections defined using ZIKV IgG3 across both DENV-naïve and DENV-exposed populations in Brazil (Figure 3). ZIKV seroconverters consistently showed higher MSD NS1 responses compared with non-seroconverters, while DENV NS1 signals remained stable.

**Figure 3.**
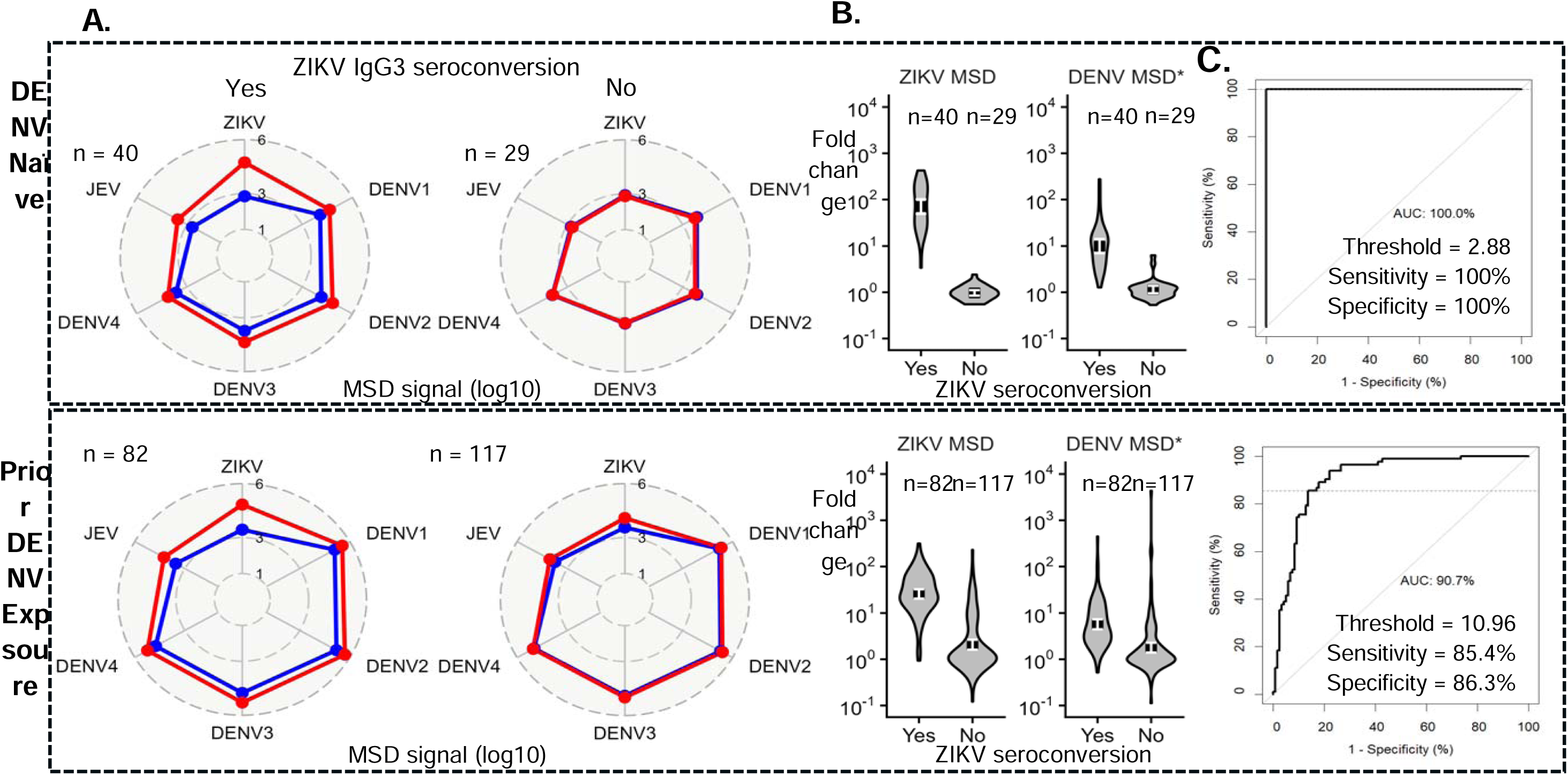
Application of the MSD assay to seroincidence studies. (A) Radar plots present MSD signal intensities (log₁₀) for ZIKV, DENV1–4, and JEV antigens among participants with (red) and without (blue) ZIKV IgG3 seroconversion, stratified by DENV exposure status (top: DENV-naïve; bottom: DENV-exposed). (B) Violin plots display fold changes in MSD signals for ZIKV and DENV antigens according to ZIKV IgG3 seroconversion status. Participants with ZIKV seroconversion exhibited higher fold changes compared to non-seroconverters. (C) ROC curve analyses identify the optimal fold change thresholds to classify ZIKV seroconversion: 2.88 for DENV-naïve (AUC = 100%) and 10.96 for prior DENV-exposed participants (AUC = 90.7%).

ROC analyses revealed excellent diagnostic performance. Among DENV-naïve participants, MSD ZIKV NS1 fold-changes achieved complete separation between infected and uninfected individuals (AUC = 100%, optimal threshold = 758.58, sensitivity = 100%, specificity = 100%). Large variation in the optimal threshold was found using 10-fold cross validation (optimal threshold = 2.29-33.88) due to the considerable separation in antibody response between infected and uninfected (Table S2, sensitivity = 100-100%, specificity = 100-100%). Among participants with prior DENV exposure, assay performance remained strong (AUC = 90.7%, sensitivity = 85.4%, specificity = 86.3%), though the optimal discrimination threshold shifted to fold-change of 10.96, reflecting the influence of pre-existing flavivirus immunity on background antibody and response levels. Variation in the optimal threshold was lower for participants with prior DENV exposure using the 10-fold CV approach (optimal threshold = 2.88-25.70), with a wider range of testing performance metrics (Table S2, sensitivity = 75-100%, specificity = 58-100%).

The threshold derived from DENV-exposed participants, fold-change ≥ 10.96 was selected as the standardized cutoff to maximize the performance of the ZIKV infection definition, ensuring high specificity while maintaining sensitivity across heterogeneous immune backgrounds. This definition, paired with the comparison of ZIKV and DENV ratios, was subsequently applied in all cohort-based seroincidence analyses. For the PRNT confirmed ZIKV infections this threshold produces an AUC of 100% and 85.8% in DENV-naïve and -exposed participants respectively. In an independent analysis trained on these intervals with available PRNT data (n=117), the optimal threshold for the ZIKV MSD NS1 response was defined as 6.17 for classification of ZIKV infections (range of 1.86 – 6.61, sensitivity = 92-100%, specificity = 91-98%), consistent with the range obtained by cross-validation under the IgG3 serological definition.

We then assessed the sensitivity and specificity of the MSD based dual-threshold approach against virologically confirmed infections. Among PCR-confirmed DENV infections in the Thai cohort (n=12), the dual-thresholding approach improved specificity from 50% (6/12) to 83.3% (10/12). Both remaining false positives occurred in samples taken two to four weeks after a PCR-confirmed DENV infection, when DENV NS1 responses are near peak. Sensitivity could not be accurately assessed in the symptomatic patient cohorts since neither include pre-infection sera, with first samples frequently collected after symptom onset, leading to attenuated fold-changes driven by sample timing rather than assay performance.

### Household-level seroconversion dynamics in Thailand (Figure 4)

Figure 4 illustrates the serological trajectories of three participants (A–C) from a single household in the Thai cohort. The figure is composed of three panels: the first panel shows the MSD platform signal for antigens of Zika virus (ZIKV) and Dengue virus serotypes 1–4 (DENV1-4); the second panel illustrates the fold-change in ZIKV antibody signal over time; and the rightmost panel depicts the ratio of ZIKV fold-change to the maximum DENV fold-change (ZIKV/DENV). A shaded grey interval marks the follow-up period during which all participants satisfied both predefined criteria: (1) ZIKV fold-change ≥ 10.96 and (2) ZIKV/DENV ratio > 1. In the MSD signal panel, a black arrow indicates the time point at which participants B and C tested positive for DENV-1 by PCR.

**Figure 4.**
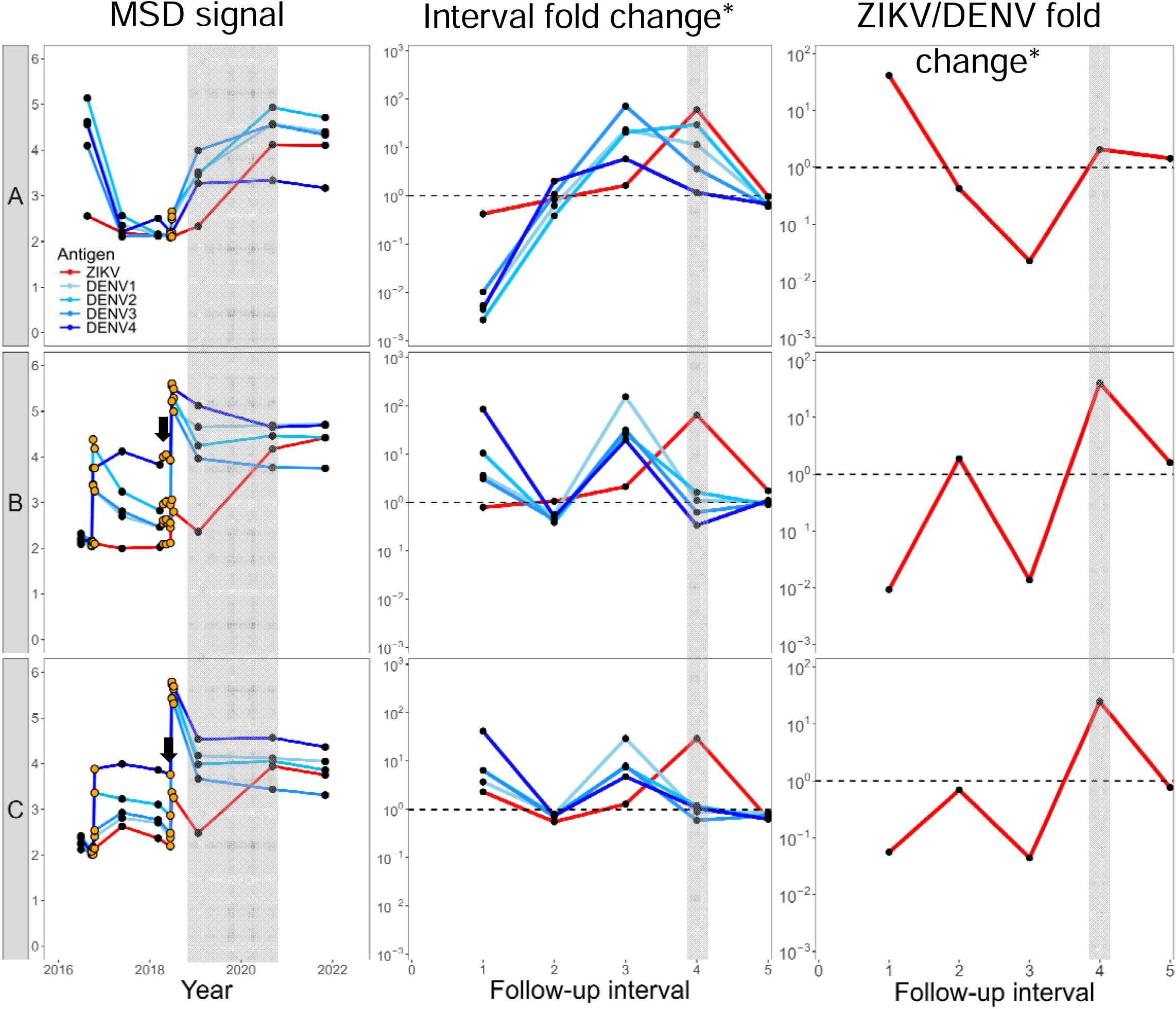
Example of seroconversion over time within a household from the Thai cohort. Longitudinal antibody responses are shown for three household participants aged 0, 6, and 12 at enrollment (A–C). Each panel illustrates (i) MSD signal intensity (log₁₀) for ZIKV and DENV1–4, (ii) interval fold changes in MSD signals, and (iii) ZIKV/DENV fold change ratios. Red lines indicate ZIKV responses, while blue lines indicate DENV serotypes. Black points represent yearly interval data, orange points represent acute and convalescent data, and the black arrow defines a PCR confirmed DENV infection event. Shaded areas represent the follow-up interval where seroconversion events were detected.

In the MSD signal panel, participants B and C show a pronounced jump in signal shortly after the PCR-confirmed DENV-1 infection (black arrow) during the third follow-up interval that triggered acute and convalescent sampling across the household (orange points). During this same interval participant A displayed elevations in their DENV signal that were sufficiently high to be indicative of an infection event but had no reported symptomatic events. The high DENV fold change and its relative magnitude to the ZIKV fold change within this third interval can be seen in the second and third panels respectively for each participant.

In the grey-shaded interval (interval 4), all three participants crossed the ≥ 10.96 ratio threshold for ZIKV (interval 4), consistent with our first criterion for seroconversion. Temporal alignment suggests that this seroconversion occurred between 2019 and 2020. In the ZIKV/DENV ratio panel, all participants reached values > 1 during the same interval, thereby fulfilling the second criterion that the ZIKV fold-change exceeded the maximal DENV fold-change in that period.

Together, these results suggest that all household members experienced seroconversion of ZIKV within the same interval (interval four), meeting both serological criteria concurrently. The synchronicity of these events within a single household is consistent with possible intra-household transmission of ZIKV.

Use of the fold-change threshold alone yielded a specificity of 74%, reflecting the well-recognized challenge of distinguishing ZIKV responses from DENV-induced boosting in DENV endemic settings. The incorporation of the ZIKV/DENV ratio panel substantially improved specificity to 92.5%, demonstrating that the dual-threshold framework mitigates cross-reactive misclassification.

Together, these results indicate that all three household members met the serological criteria for ZIKV seroconversion within the same interval (2019–2020). The spatiotemporal clustering of seroconversion within a single household is suggestive of intra-household ZIKV transmission, although asynchronous community-acquired infections occurring within the yearlong sampling interval cannot be excluded.

### Visualization of confirmed DENV infections and ZIKV seroconversions in reduced dimensions (Figure 5)

Figure 5 presents paired serological samples visualized in two-dimensional space using PCA. Panel A shows PCR-confirmed Dengue virus (DENV) and Zika virus (ZIKV) seroconversions from Brazil, while panels B and C show the remaining paired samples from Thailand and Brazil, respectively. All samples are colored according to the ZIKV infection discrimination criteria defined above. Figure 5 includes ZIKV seroconversions from the Brazil community cohort confirmed using IgG3 seroconversion, providing a complementary validation approach to the PCR-confirmed infections shown in Figure 1.

**Figure 5.** Dimension reduction of interval ratios. Principal component analysis (PCA) of NS1-specific IgG responses to six flavivirus antigens (DENV-1–4, JEV, and ZIKV) measured by the Mesoscale Discovery System. Arrows represent variable loadings: blue arrows indicate DENV serotype-specific responses, red arrows represent ZIKV, and green arrows denote JEV. (A) PCA projection of Brazilian samples with laboratory-confirmed ZIKV infections (blue squares, NS1 IgG3-confirmed) and Thai samples with laboratory-confirmed DENV infections (red circles, PCR-confirmed). (B–C) PCA projections of Thai (B) and Brazilian (C) samples classified by ZIKV fold-change (FC) thresholds. Blue squares indicate ZIKV FC ≥ 10.96 and greater than DENV FC, yellow circles indicate ZIKV FC ≥ 10.96 only, and gray triangles represent ZIKV FC < 10.96.

We performed PCA on the ratio of MSD signal between paired samples across all four DENV serotypes, Japanese encephalitis virus (JEV), and ZIKV. The vector components representing each of these six ratios used to construct the first two dimensions are colored in red, green, and blue. In all panels, the centroid for each group is presented as a larger version of the corresponding color and shape combination. The first two dimensions account for 93.1% of the variation in the data.

Panel A presents RT-PCR-confirmed DENV infections from Thailand (red circles) and suspected ZIKV seroconversions from Brazil defined using ZIKV IgG3 (dark blue rectangles). The JEV and ZIKV ratio vectors both point toward the cluster of ZIKV seroconversions, while all four DENV serotype vectors point toward the confirmed DENV infections. This demonstrates that infections are differentiated along the DENV and the ZIKV/JEV axes, although a few ZIKV seroconversions cluster among the DENV infections, likely due to cross-reactivity.

Panels B and C subset the data to samples from the Thai and Brazilian cohorts respectively, with points colored by the dual-thresholding approach: yellow indicates ZIKV fold change ≥ 10.96, and light blue indicates ZIKV fold change > DENV fold change. Samples without a ZIKV fold change increase ≥ 10.96 are colored gray. Samples where ZIKV fold change is ≥ 10.96 but ≤ DENV fold change occupy similar space to confirmed DENV infections, suggesting these represent cross-reactivity rather than true ZIKV infections. Incorporating this dual-thresholding approach increases overlap between inferred ZIKV infections and the confirmed ZIKV seroconversions, thereby reducing false positive classifications compared to using ZIKV fold change alone.

### Frequency of ZIKV infections by cohort and case definition (Table 1)

Using the MSD multiplex electrochemiluminescence assay, we estimated the frequency of ZIKV infections across multiple cohorts by applying two case definitions based on fold-change (FC) in ZIKV NS1 responses. The first definition classified participants as positive when ZIKV FC ≥ 10.96, while the second required both ZIKV FC ≥ 10.96 and > maximum DENV (1–4) FC within the same individual. These complementary thresholds were used to evaluate the impact of pre-existing DENV immunity on the serological discrimination of ZIKV infection. A summary of these results, including cohort sizes, follow-up durations, infection frequencies, and event rates, is presented in Table 1.

**Table 1.**
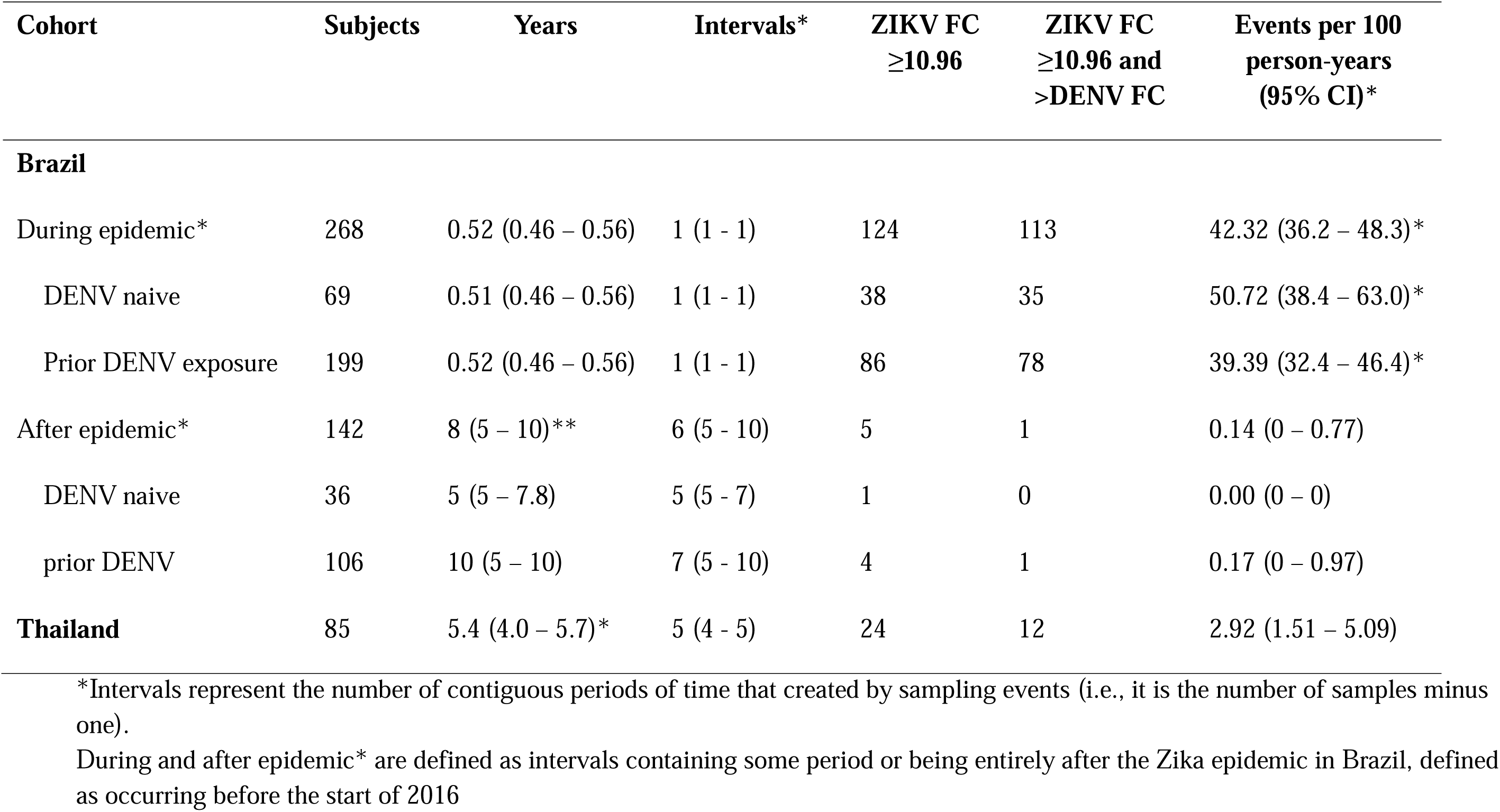
Attack Rate of ZIKV Serologic Response in Brazil and Thailand Cohorts.

Application of the dual-threshold framework to the Brazil cohort revealed distinct transmission patterns across epidemic and post-epidemic periods. During the 2015–2016 epidemic, we observed high ZIKV incidence, consistent with widespread transmission during this period. In contrast, the post-epidemic period (2017–2024) showed very low transmission, with incidence rates decreasing substantially. The ability to detect these temporal patterns demonstrates the utility of the MSD assay for monitoring ZIKV transmission dynamics over time.

During the 2015–2016 epidemic in Brazil, 124 of 268 participants (46.3%) met the ZIKV FC ≥ 10.96 criterion, while 113 (42.1%) met the stricter combined definition, corresponding to an incidence rate of 42.32 (95% CI: 36.2 – 48.3) events per 100 person-years. Among DENV-naïve participants, infection frequency was slightly higher (38/69 vs 35/69; 55.1% vs 50.7% for the two definitions), yielding an event rate of 50.72 (38.4 – 63.0) per 100 person-years. For participants with prior DENV exposure, 43.2% met the primary definition and 39.2% the relative definition, corresponding to 39.39 (32.4 – 46.4) events per 100 person-years.

In the post-epidemic follow-up period (2017–2024), only 5 of 142 participants (3.5%) showed a ZIKV FC ≥ 10.96, and just 1 (0.7%) met the combined definition, with the overall event rate decreasing to 0.14 (0 – 0.77) per 100 person-years. No ZIKV seroconversions were detected among DENV-naïve individuals, and only one seroconversion was observed among those with prior DENV exposure (0.17 [0 – 0.97] per 100 person-years).

Consistency of the dual-thresholding approach with PRNT and IgG3 inference approaches in Brazil were estimated by comparing the incidence rate for intervals where all three data streams were available. We find that in this subset (117 subjects), contained entirely during the ZIKV epidemic, all three ZIKV infection inference approaches are consistent with one another and report higher incidence rates per 100 person-years (PRNT: 95.4, 95% CI: 77.3 – 113.5; IgG3: 89.8, 95% CI: 71.6 – 108.1; ZIKV FC: 82.9, 95% CI: 64.9 – 101.0) than the full dataset analyzed here.

In the Thailand cohort, 24 of 85 participants (28.2%) met the primary ZIKV FC ≥ 10.96 criterion, and 12 (14.1%) met the stricter one, corresponding to an event rate of 2.92 (1.51 – 5.09) per 100 person-years.

## Discussion

We validated and applied a multiplex electrochemiluminescence assay targeting NS1 antigens of Zika virus (ZIKV) and dengue virus (DENV) across populations with distinct flavivirus immune backgrounds. Four principal findings emerge from this study. First, the MSD ZIKV NS1 assay generated strong and sustained antibody responses following PCR-confirmed ZIKV infection. Second, the assay showed minimal cross-reactivity following primary DENV infection and only transient low-level reactivity after post-primary DENV infections. Third, MSD signal intensities correlated strongly with PRNT_50_ neutralization titres for each virus (ZIKV and DENV), supporting the assay as a quantitative surrogate for functional serological assays, while remaining minimally cross-reactive between viruses. Fourth, we identified and validated a dual-threshold framework based on a ZIKV fold-change of at least 10·96 combined with a ZIKV-to-DENV fold-change ratio greater than 1, which substantially improved specificity for identifying ZIKV infection in DENV-endemic settings. Together, these findings support the use of multiplex NS1 electrochemiluminescence assays for serological discrimination, seroincidence estimation, and transmission inference in flavivirus co-endemic regions.

The identification of a standardized seroconversion threshold is particularly relevant for large-scale epidemiological studies. Receiver operating characteristic analyses demonstrated excellent discrimination between ZIKV seroconverters and non-seroconverters, including among participants with previous DENV exposure. Although NS1-based assays have previously shown improved specificity compared with envelope-based serology, interpretation of flavivirus antibody responses remains challenging in populations with repeated DENV exposure because anamnestic immune responses can generate substantial cross-reactivity. Our second criterion, the requirement that the ZIKV fold-change exceeds the maximum DENV fold-change within the same individual—was designed to address this limitation directly. This refinement substantially reduced likely false-positive classifications attributable to DENV boosting while maintaining high sensitivity. The marked reduction in inferred ZIKV infections in Thailand after application of the second criterion illustrates the importance of accounting for heterogeneous flavivirus immune histories when interpreting serological data in hyperendemic settings.

Application of this framework to longitudinal cohorts generated epidemiologically coherent transmission patterns. In Brazil, we observed high ZIKV seroincidence during the 2015–16 epidemic followed by very low post-epidemic transmission through 2024, consistent with rapid accumulation of population immunity after explosive emergence ^27,37,38^.

The relatively modest reduction in case classification after application of the second criterion in Brazil during the epidemic suggests that cross-reactive misclassification was limited during these years, however more modern samples may provide higher cross-reactive profiles as demonstrated by the substantive decline in the albeit small number of identified infections after the ZIKV epidemic (from 5 to 1). In Thailand we see the impact of this highly endemic DENV setting, concurrently high JEV vaccination rate, presenting a difficult setting where cross-reactivity-driven false positives account for 50% of identified ZIKV infection, demonstrating the immunological complexity that these related flaviviruses create and suggest the value of the dual-threshold approach for improving specificity. These findings support the broader use of longitudinal quantitative serological frameworks to reconstruct transmission dynamics where routine virological surveillance is limited or absent.

Our household-level analyses further illustrate the epidemiological utility of this approach. The temporal clustering of seroconversion among members of a single Thai household suggests possible intra-household ZIKV transmission. Although the yearly sampling interval precludes definitive reconstruction of transmission chains and does not exclude asynchronous community-acquired infections, these findings demonstrate how quantitative longitudinal serology may support transmission inference in settings where direct virological confirmation is difficult to obtain. Integration of serological data with entomological, genomic, and spatial epidemiological information could further strengthen these approaches for outbreak investigation and arbovirus surveillance.

The strong correlations between MSD NS1 signals and PRNT_50_ titres across ZIKV and all four DENV serotypes support the biological validity of the assay and suggest that multiplex electrochemiluminescence platforms could provide scalable alternatives to labor-intensive neutralization assays. This scalability is important because PRNT-based approaches are often impractical for large cohort studies, biobank characterization, or long-term population surveillance. By enabling simultaneous quantification of antibody responses against multiple flaviviruses, the MSD platform may help address a longstanding limitation in arbovirus epidemiology: the lack of high-throughput tools capable of reliably distinguishing closely related flavivirus infections in endemic populations. ^19–21^

These findings also have implications beyond surveillance. In regions where DENV and ZIKV co-circulate, interpretation of vaccine-induced immune responses remains complicated by extensive serological cross-reactivity. A scalable assay that quantitatively correlates with neutralization titres could support vaccine evaluation studies, facilitate identification of immune correlates of protection, and improve interpretation of longitudinal immunity following natural infection or vaccination. In addition, the ability to identify low-level or silent ZIKV circulation after epidemic periods may strengthen preparedness strategies for future outbreaks and support more targeted public health interventions.

This study has several limitations. First, the dual-threshold framework performed consistently across the cohorts included here, but validation in additional epidemiological settings with distinct flavivirus exposure histories will be necessary before broad implementation. Second, the ratio-based criterion requires longitudinal paired samples with comparable sampling intervals and concurrent DENV measurements, which may limit applicability in settings with irregular follow-up or incomplete sampling. Third, although the household clustering findings are suggestive of intra-household transmission, we lacked virological and entomological data to confirm direct transmission pathways. Finally, because flavivirus immunity is shaped by complex sequential exposures, additional studies will be needed to determine how prior exposure to other flaviviruses, including yellow fever virus and Japanese encephalitis virus, influences assay performance.

In conclusion, the dual-threshold serological framework presented here addresses a major barrier in flavivirus epidemiology: the reliable differentiation of ZIKV and DENV infections at population scale. By combining high-throughput multiplex electrochemiluminescence technology with quantitative longitudinal discrimination criteria validated across multiple cohorts and immune backgrounds, this approach enables scalable seroincidence estimation that would otherwise require resource-intensive neutralization assays. As ZIKV continues to circulate at low levels with potential for re-emergence, accurate serological discrimination will remain essential for surveillance, vaccine evaluation, and transmission studies in flavivirus-endemic regions.

## Data Availability

De-identified data underlying the analyses are available upon reasonable request to the corresponding author, subject to the data-sharing agreements and ethical approvals governing the contributing cohorts.

## Disclaimer

The views, opinions and/or findings contained in this presentation are those of the author and do not necessarily reflect the views of the Department of War and should not be construed as an official DoW/Army position, policy or decision unless so designated by other documentation. No official endorsement should be made. Reference herein to any specific commercial products, process, or service by trade name, trademark, manufacturer, or otherwise, does not necessarily constitute or imply its endorsement, recommendation, or favoring by the U.S. Government.\

