## Supplementary Tables S1-S2 and Figures S1-S6 for "Applying a multiplex electrochemiluminescence NS1 assay to detect and differentiate Zika and dengue virus exposures in long-term community cohorts in Brazil and Thailand"

**Supplemental Materials**

**Table S1. Demographic Characteristics of Patient and Community Cohorts**

| **Cohorts** | **% or median (IQR)** | **Reference** |
|---|---|---|
| **Patients** | | |
| Zika cases from Brazil (n=19) | | |
| Age (years) | 41 (21, 48) | [29] |
| Female Sex | 12 (63%) | |
| Follow-up intervals (duration, years) | 0.0 (0.0, 0.2) | |
| Zika cases in US travelers (n=7) | | |
| Age (years) | NA (NA, NA) | [30] |
| Female Sex | NA (NA%) | |
| Follow-up intervals (duration, years) | NA (NA, NA) | |
| DHIM participants (n=14) | | |
| Age (years) | 34.5 (28.8, 36.3) | [32–34] |
| Female Sex | 6 (43%) | |
| Follow-up intervals (duration, years) | 0.25 (0.08, 0.5) | |
| **Community-based** | | |
| Pau da Lima, Brazil (n=268) | | |
| Age (years) | 17 (11, 32) | [27] |
| Female Sex | 163 (61%) | |
| Follow-up intervals (duration, years) | 4.5 (0.5, 8.7) | |
| Kamphaeng Phet, Thailand (n=85) | | |
| Age (years) | 12 (1, 30) | [28] |
| Female Sex | 44 (52%) | |
| Follow-up intervals (duration, years) | 5.4 (4.0, 5.7) | |

**Table S2. Out of sample sensitivity and specificity of 10-fold cross validation for optimal thresholds across DENV-naïve and -exposed populations**

| Population | Cross-validation | Optimal-threshold | Sensitivity | Specificity |
|---|---|---|---|---|
| DENV-naïve | 1 | 2.29 | 100 | 100 |
| | 2 | 8.91 | 100 | 100 |
| | 3 | 1.78 | 100 | 100 |
| | 4 | 3.39 | 100 | 100 |
| | 5 | 1.35 | 100 | 100 |
| | 6 | 2.19 | 100 | 100 |
| | 7 | 2.04 | 100 | 100 |
| | 8 | 11.48 | 100 | 100 |
| | 9 | 6.03 | 100 | 100 |
| | 10 | 8.71 | 100 | 100 |
| DENV-exposed | 1 | 14.79 | 100 | 100 |
| | 2 | 5.13 | 100 | 91.7 |
| | 3 | 11.75 | 100 | 91.7 |
| | 4 | 8.32 | 75 | 90.9 |
| | 5 | 8.13 | 100 | 100 |
| | 6 | 11.48 | 75 | 91. 7 |
| | 7 | 2.82 | 100 | 72.7 |
| | 8 | 3.89 | 100 | 58. 3 |
| | 9 | 25.70 | 87.5 | 83. 3 |
| | 10 | 13.80 | 87.5 | 83.3 |

**Figure S1. Dynamics of antibody responses following ZIKV infection across six NS1 antigens measured by MSD assay**

Longitudinal dynamics of antibody responses to ZIKV, DENV1–4, and JEV NS1 antigens measured by the MSD multiplex electrochemiluminescence assay in individuals with confirmed ZIKV infection. Data are presented for both Brazilian subjects and U.S. travelers, corresponding to the cohorts shown in Figure 1.

Antibody levels (y-axis, log scale) are plotted over time since infection or case identification (x-axis, in days).
• Red symbols indicate acute-phase samples.
• Blue symbols represent early convalescent samples.
• Black symbols correspond to later follow-up samples.

**Figure S2. Dynamics of antibody responses following DENV infection across six NS1 antigens measured by MSD assay**

Longitudinal dynamics of antibody responses to DENV1–4, ZIKV, and JEV NS1 antigens measured by the MSD multiplex assay in participants with confirmed DENV infection. Data include Dengue Human Infection Model (DHIM) participants with primary DENV-1, 3, and 4 infection in addition to Thai cohort participants with primary and secondary DENV infections, corresponding to the cohorts shown in Figure 1.

Antibody levels (y-axis, log scale) are plotted over time since infection (x-axis, in days).
• Red symbols indicate acute-phase samples.
• Blue symbols represent early convalescent samples.
• Black symbols correspond to later follow-up samples.

**Figure S3. Application of the MSD assay to seroincidence studies using $PRNT_{50}$ as reference for ZIKV infection**

(A) Radar plots show MSD signal intensity ($log_{10}$) for ZIKV, DENV1–4, and JEV NS1 antigens among participants (n=83) before (blue) and after (red) a ZIKV $PRNT_{50}$ seroconversion (n=15 and 36) or non-event (n=13 and 19), stratified by DENV-naïve (top, n=15 and 13) and prior DENV-exposed (bottom, n=36 and 19) cohorts.

(B) Violin plots display fold changes in MSD signals for ZIKV and DENV NS1 antigens according to ZIKV $PRNT_{50}$ seroconversion status. Participants with confirmed $PRNT_{50}$ seroconversion exhibited substantially higher ZIKV fold changes compared to non-seroconverters, while DENV fold changes remained low or unchanged.

(C) Receiver operating characteristic (ROC) curve analyses identify the optimal MSD fold-change thresholds to classify ZIKV infection based on $PRNT_{50}$ seroconversion: DENV-naïve participants (AUC = 100%) and for prior DENV-exposed participants (AUC = 85.8%).

**Figure S4A. Longitudinal antibody dynamics among participants from the Brazil cohort with ZIKV fold-change ≥ 10.96**

Longitudinal antibody responses measured by the MSD multiplex assay for a randomly selected set of participants (n = 5) from the Brazil cohort who exhibited a ZIKV NS1 fold-change ≥ 10.96, consistent with ZIKV infection.

For each participant, panels display:
(i) neutralization titres (RVP),

(ii) MSD signal intensities ($log_{10}$) for ZIKV and DENV1–4 antigens,
(iii) interval fold-change in MSD signals, and
(iv) ZIKV/DENV fold-change ratios.

Red lines indicate ZIKV responses, and blue lines indicate DENV serotypes. Shaded areas mark the follow-up interval during which ZIKV seroconversion events were detected.
These profiles illustrate clear increases in ZIKV antibody responses during the epidemic period, with limited rise in DENV signals, supporting the specificity of the MSD assay in identifying ZIKV infections within the Brazilian cohort.

**Figure S4B. Longitudinal antibody dynamics among participants from the Thailand cohort with ZIKV fold-change ≥ 10.96**

Longitudinal antibody responses measured by the MSD multiplex assay for participants (n = 24) from the Thailand cohort who demonstrated a ZIKV NS1 fold-change ≥ 10.96, meeting the threshold for ZIKV infection.

For each participant, panels display:
(i) neutralization titres (RVP),
(ii) MSD signal intensities ($log_{10}$) for ZIKV and DENV1–4 antigens,
(iii) interval fold-change in MSD signals, and
(iv) ZIKV/DENV fold-change ratios.

Red lines denote ZIKV responses and blue lines denote DENV responses. Shaded intervals highlight the follow-up period when ZIKV seroconversion occurred.
These data illustrate the temporal dynamics of antibody responses in the Thai community cohort, where multiple participants exhibited distinct ZIKV seroconversion events amid a background of pre-existing DENV immunity.

**Figure S5. Temporal trajectories of DENV and ZIKV ratios after confirmed infections.** Polymerase Chain Reaction (PCR) confirmed DENV infections and ZIKV seroconversions are represented as red and blue lines respectively. (A) The mean log10 DENV-1 to 4 interval ratios and (B) log10 ZIKV interval ratios for each individual along with a loess fit for each infection group.

**Figure S6. Correlation of MSD and IgG3 responses to ZIKV in pre- and post-outbreak samples**

Correlation between ZIKV NS1 antibody levels measured by the MSD multiplex assay and ZIKV-specific IgG3 optical density (nOD) in serum samples collected before (Pre) and after (Post) the 2015–2016 Zika virus outbreak in Salvador, Brazil. Each point represents an individual sample; the blue lines indicate linear regression fits, and red dashed lines mark the lower detection limits for each assay.

**Figure S1. Longitudinal Serologic Responses to ZIKV, DENV1-4, and JEV in ZIKV Infections cohorts**

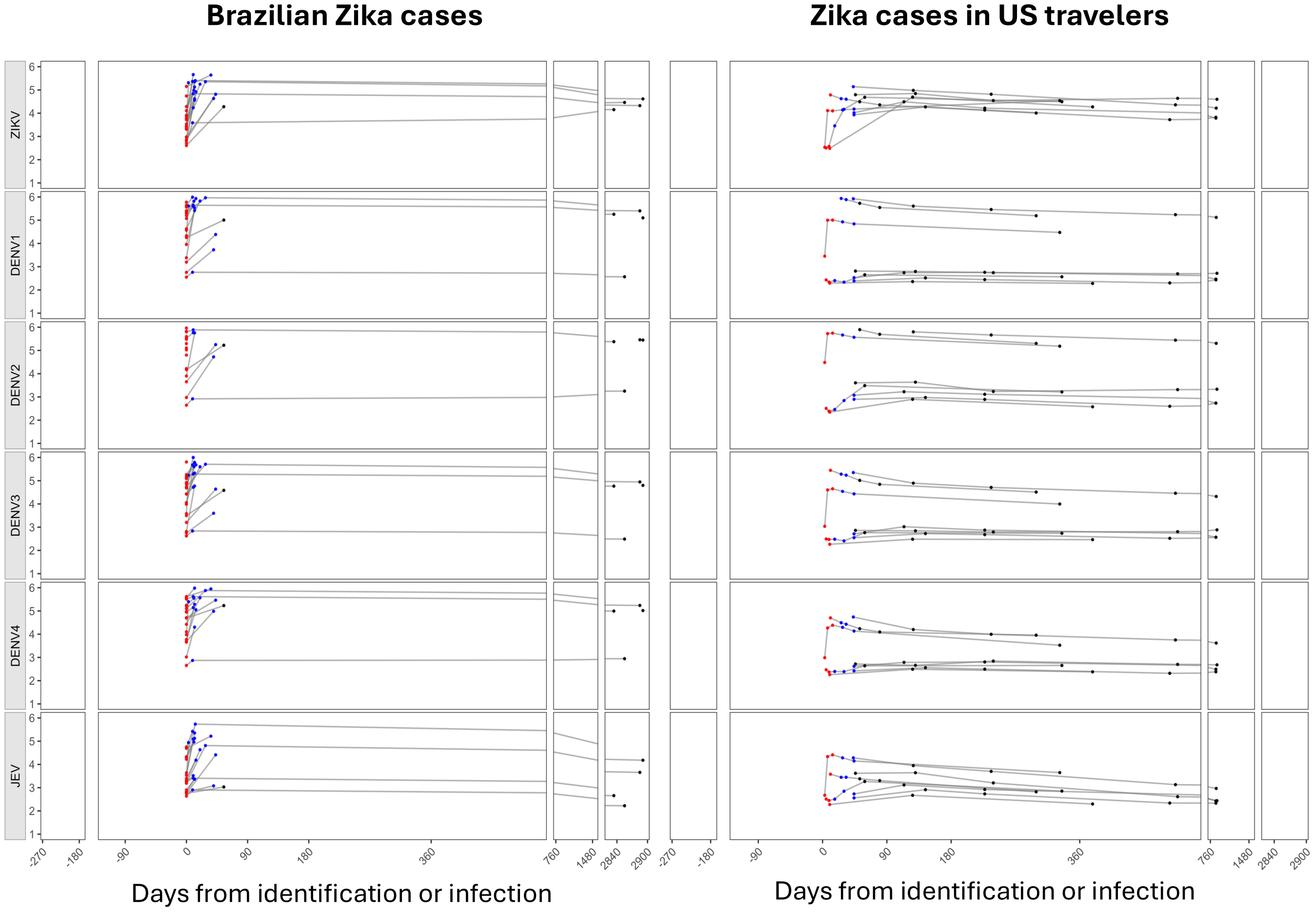

### Figure S2. Longitudinal Serologic Responses to ZIKV, DENV1-4, and JEV in DENV Infections cohorts

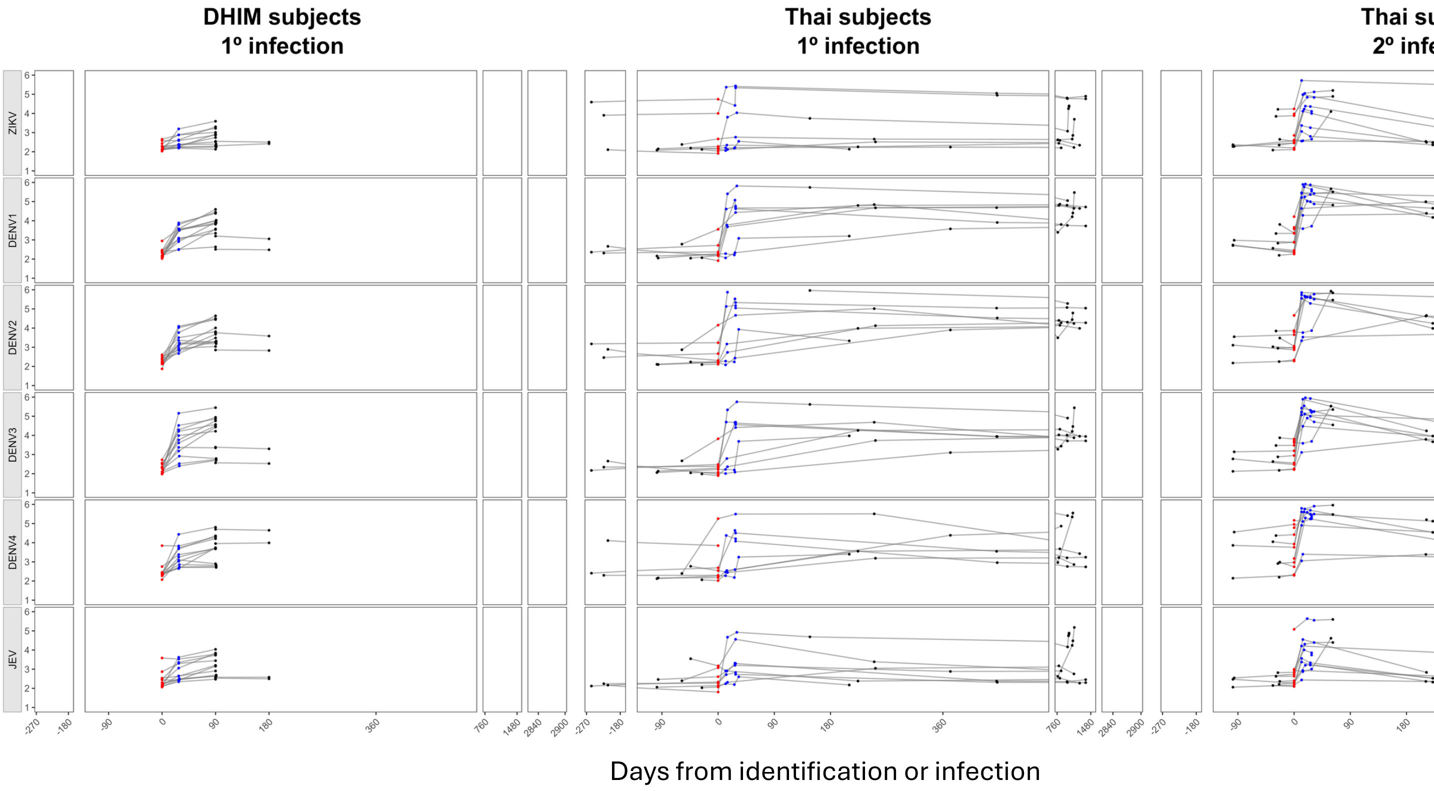

**Figure S3: Application of MSD Assay to Seroincidence Studies - PRNT$_{50}$-defined ZIKV**

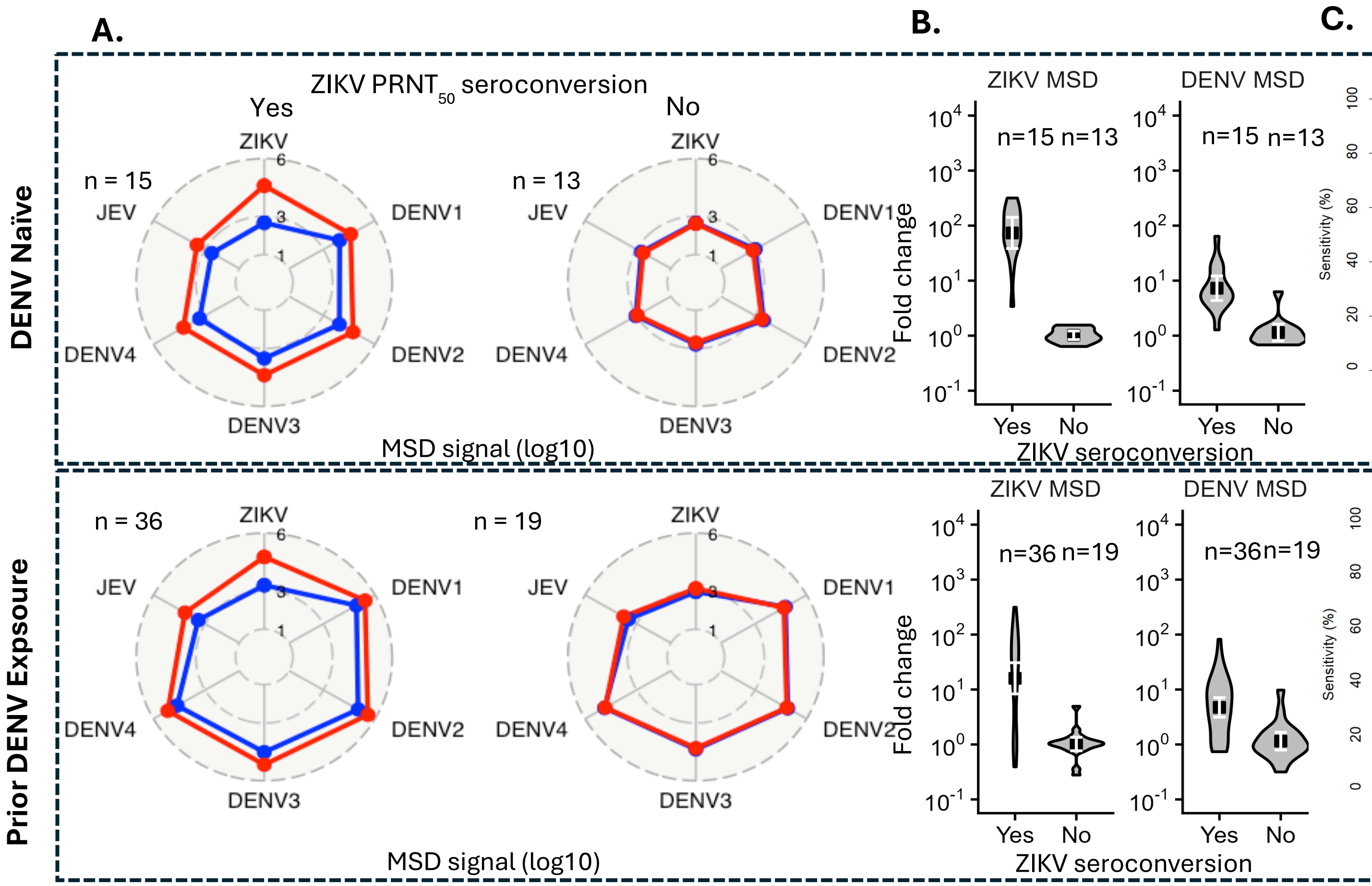

**Figure S4A. Participants (n = 5) from the Brazil cohort with fold-change ≥ 10.96.**

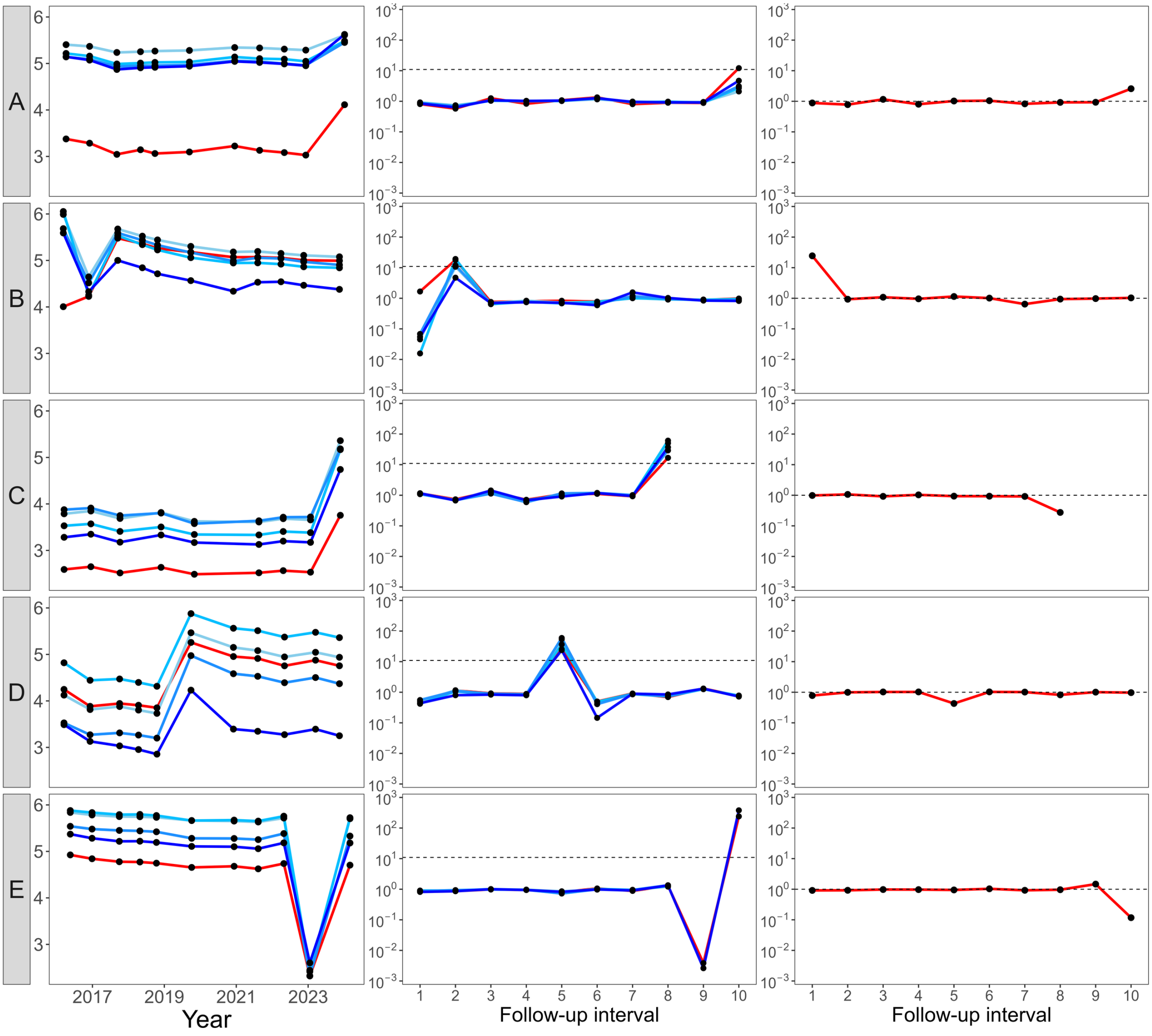

**Figure S4B. Participants (n = 24) from the Thailand cohort with fold-change ≥ 10.96.**

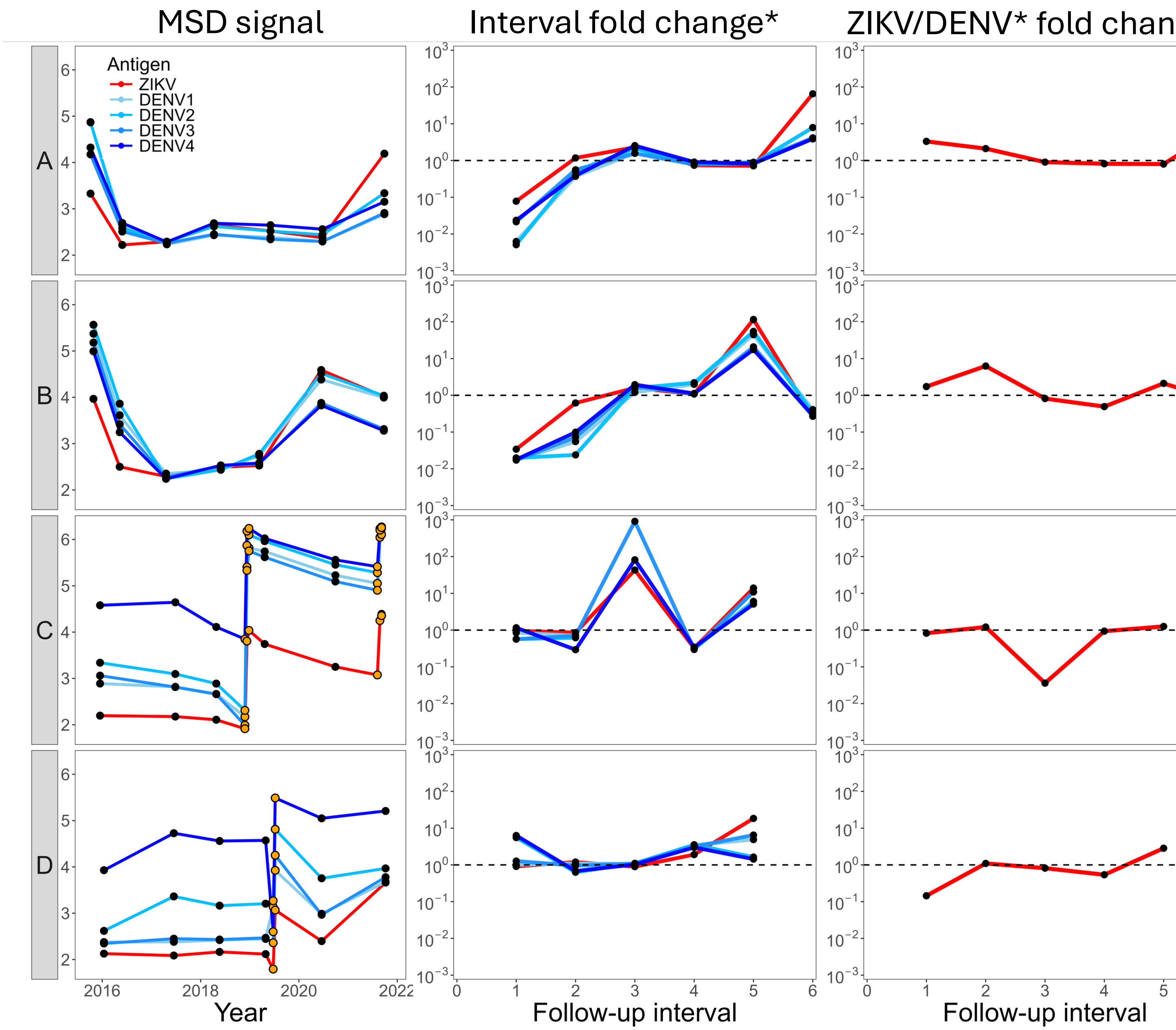

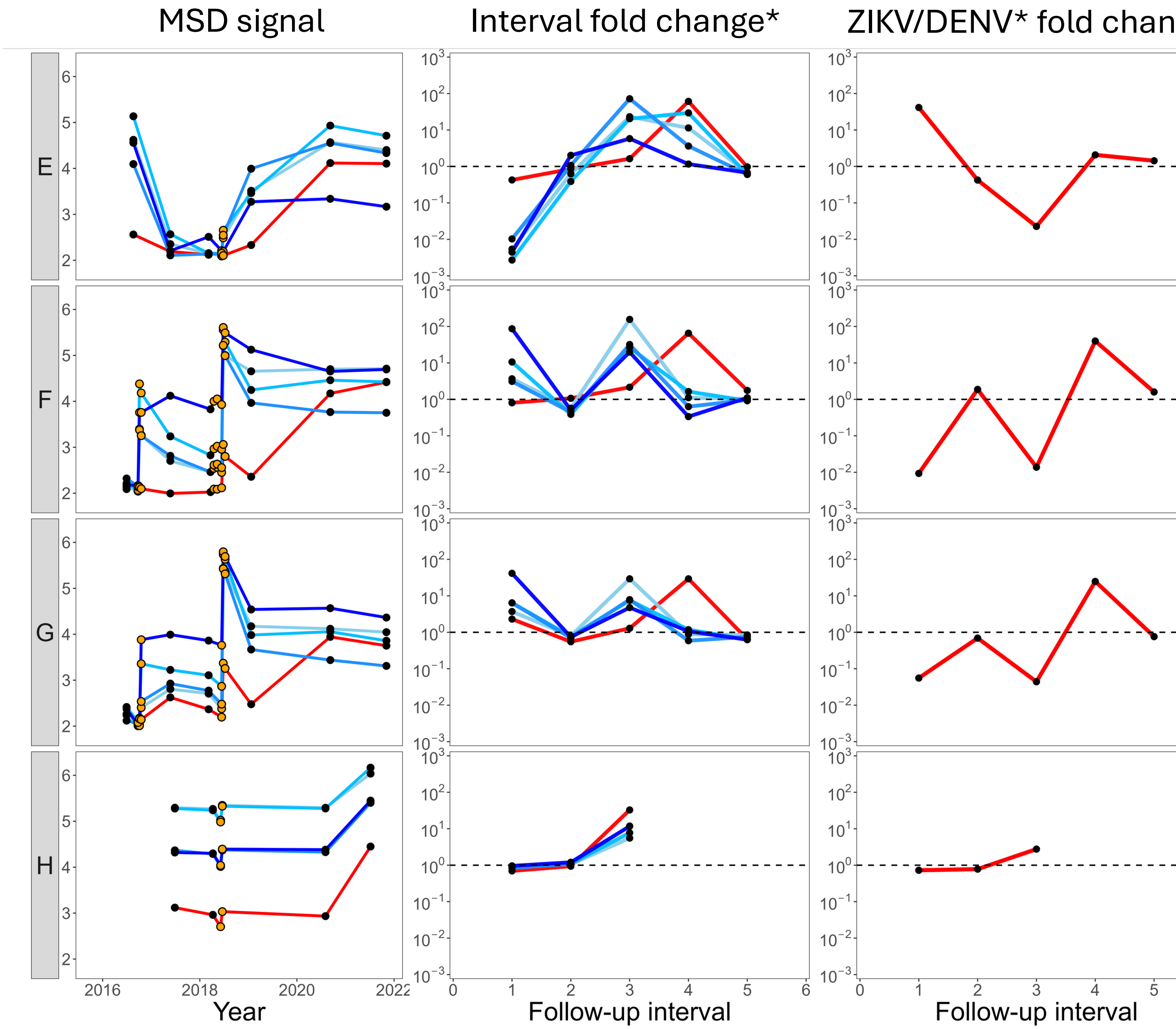

MSD signal
Interval fold change*
ZIKV/DENV* fold chan
E
F
G
H
Year
Follow-up interval
Follow-up interval

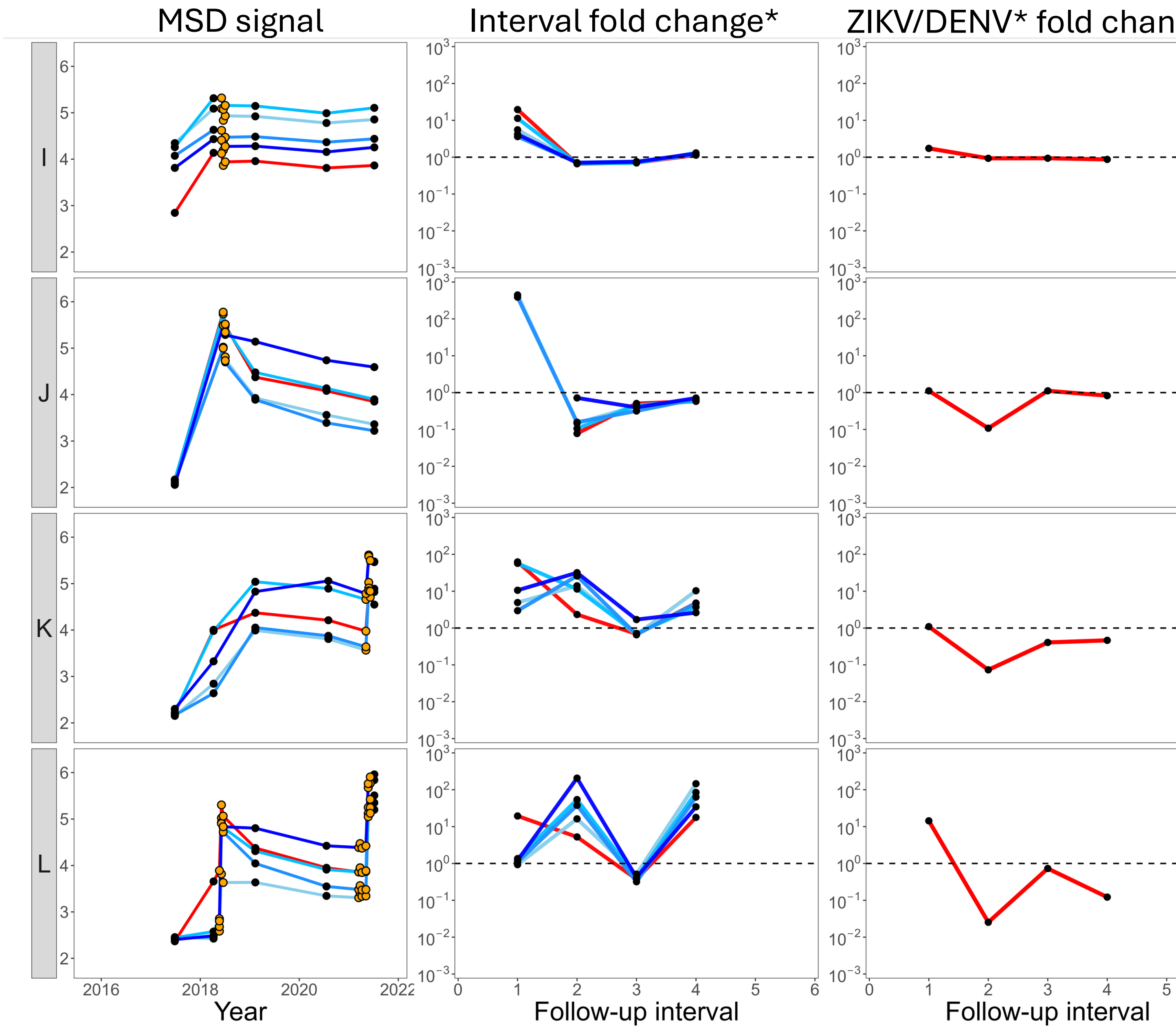

MSD signal
Interval fold change*
ZIKV/DENV* fold chan
I
J
K
L
Year
Follow-up interval
Follow-up interval
2016
2018
2020
2022

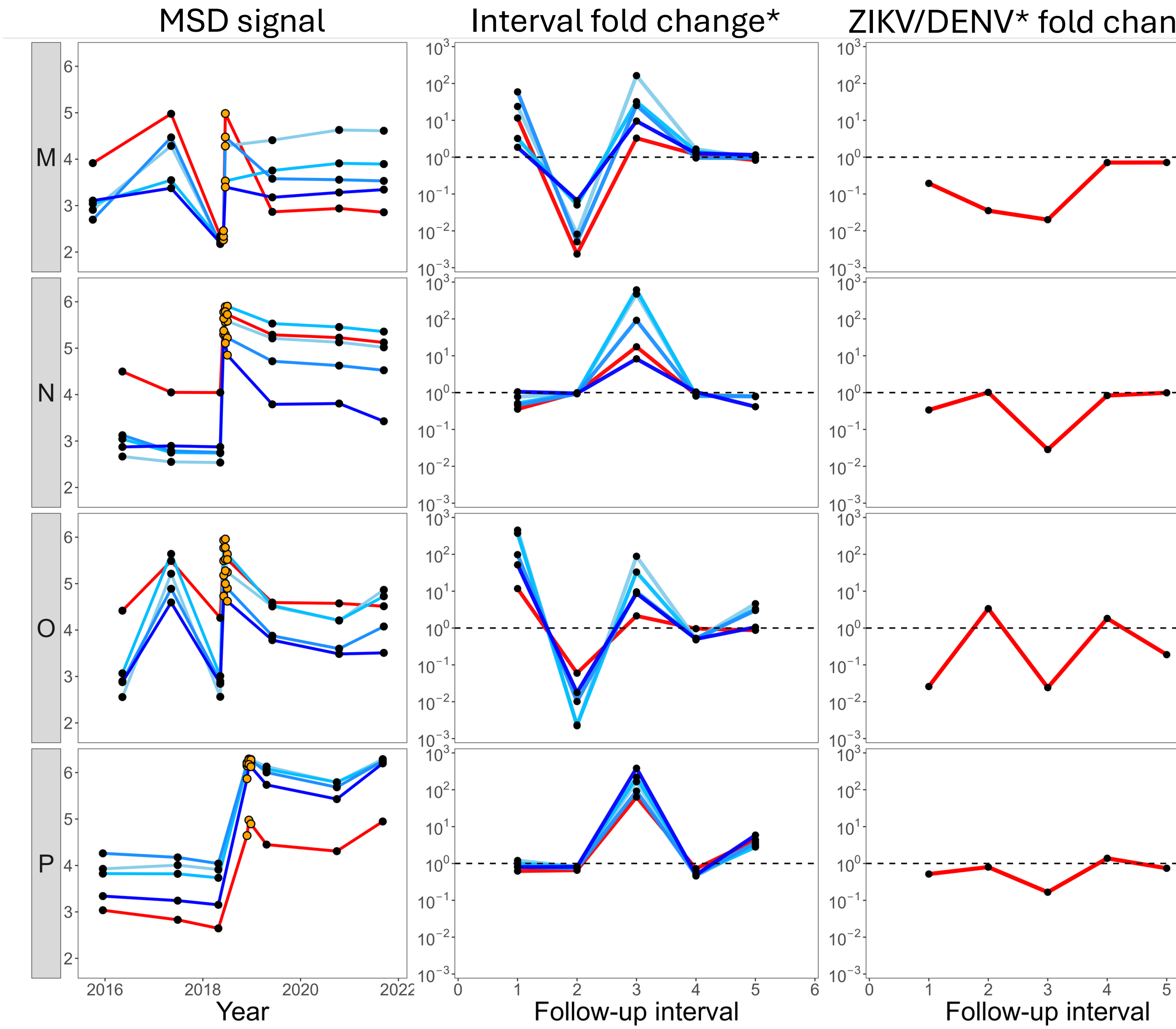

MSD signal
Interval fold change*
ZIKV/DENV* fold chan
M
N
O
P
Year
Follow-up interval
Follow-up interval

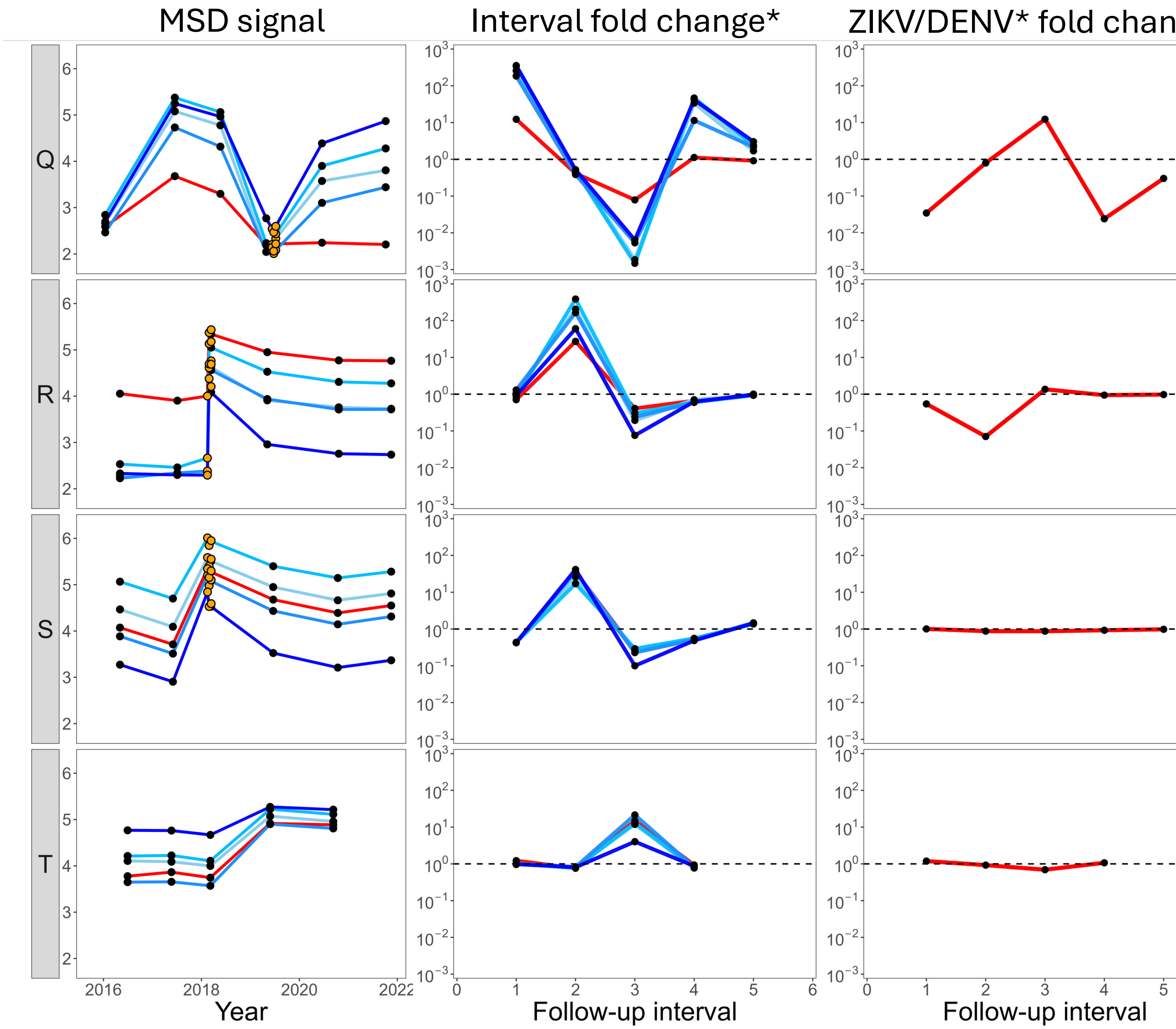
MSD signal
Interval fold change*
ZIKV/DENV* fold chan
Q
R
S
T
Year
Follow-up interval
Follow-up interval

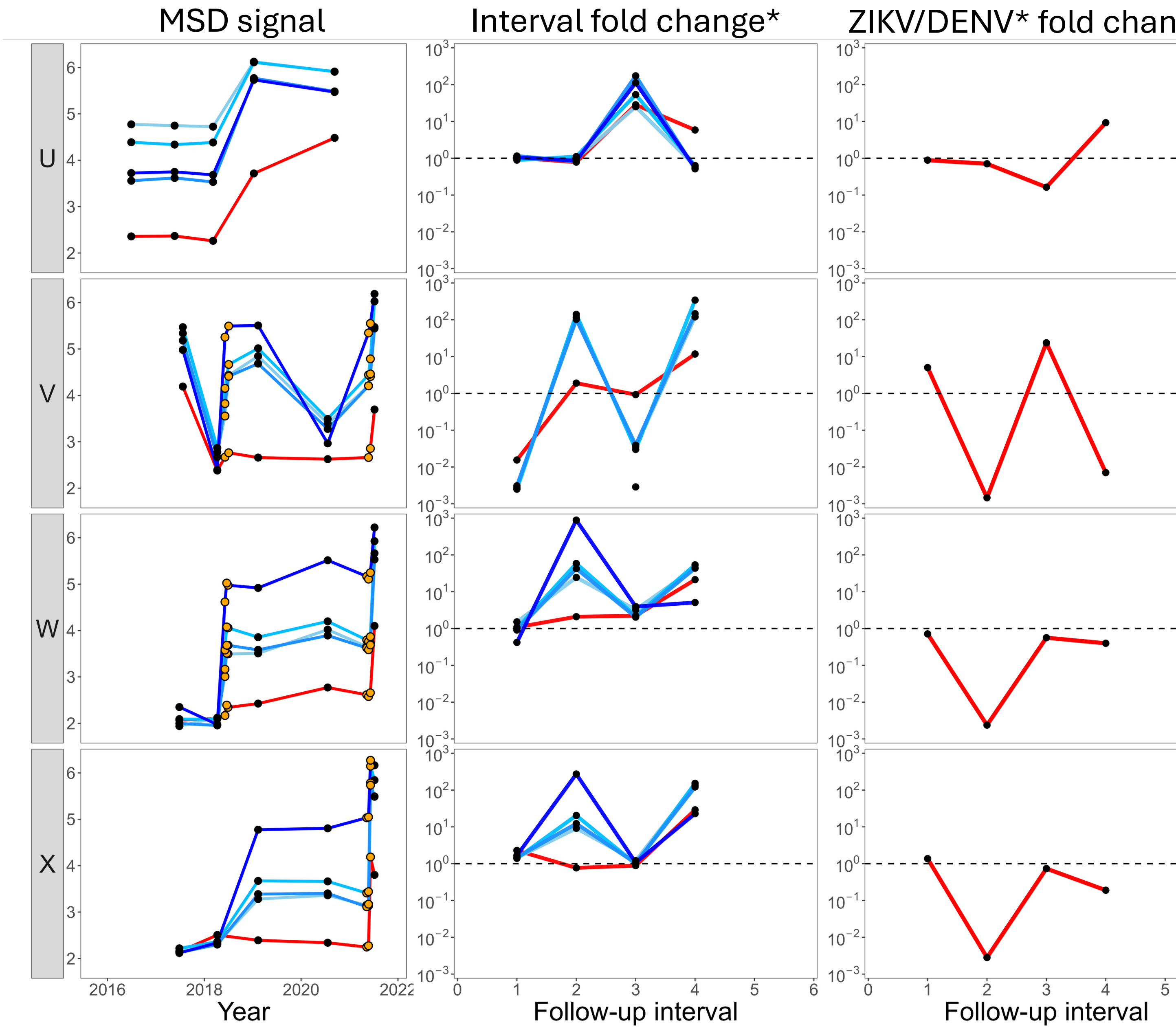

MSD signal
Interval fold change*
ZIKV/DENV* fold chan
U
V
W
X
Year
Follow-up interval
Follow-up interval
2016
2018
2020
2022

**Figure S5: Temporal trajectories of DENV and ZIKV ratios after confirmed infections.**

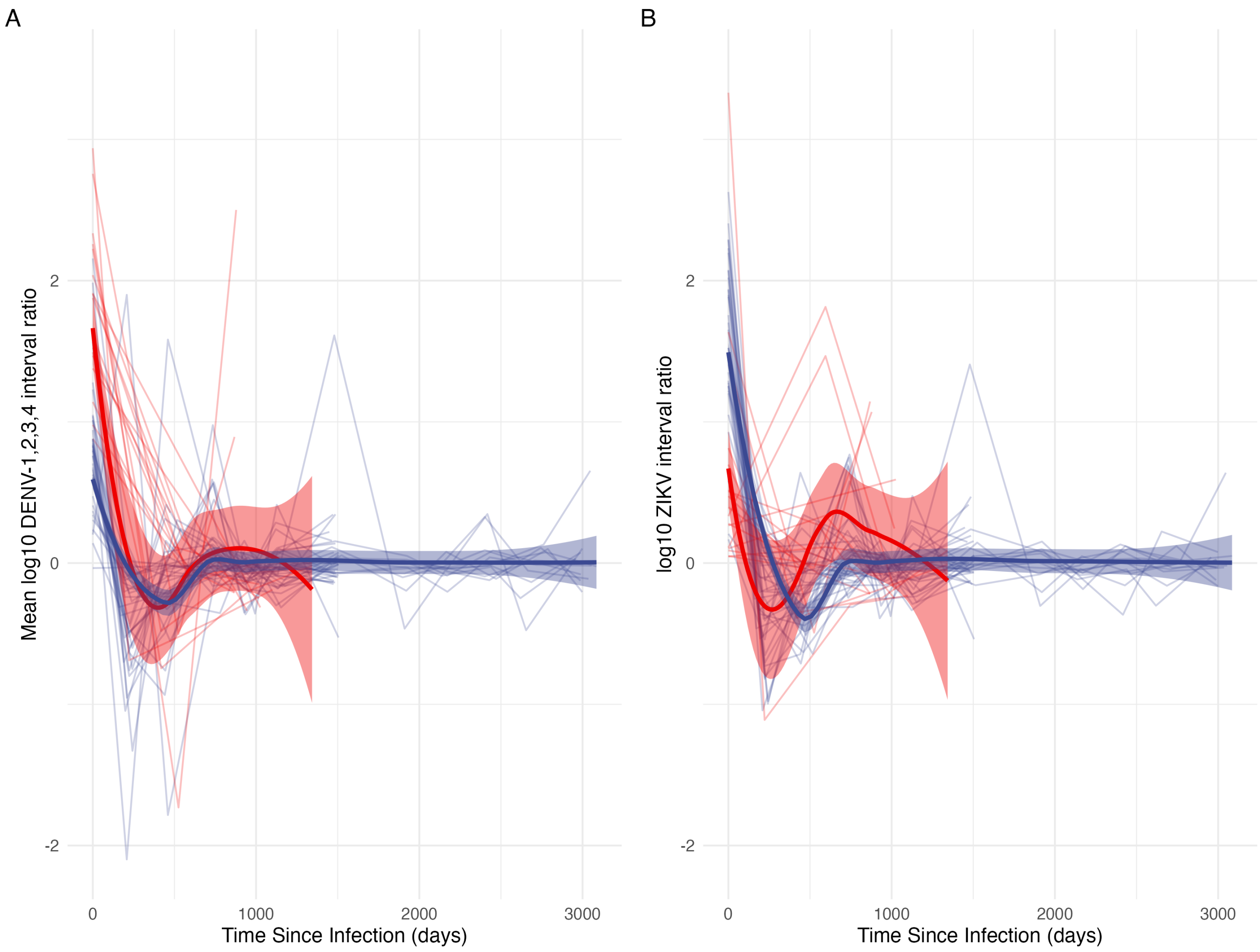


**Figure S6: Correlation of MSD and IgG3 responses to ZIKV in pre- and post-outbreak samples**

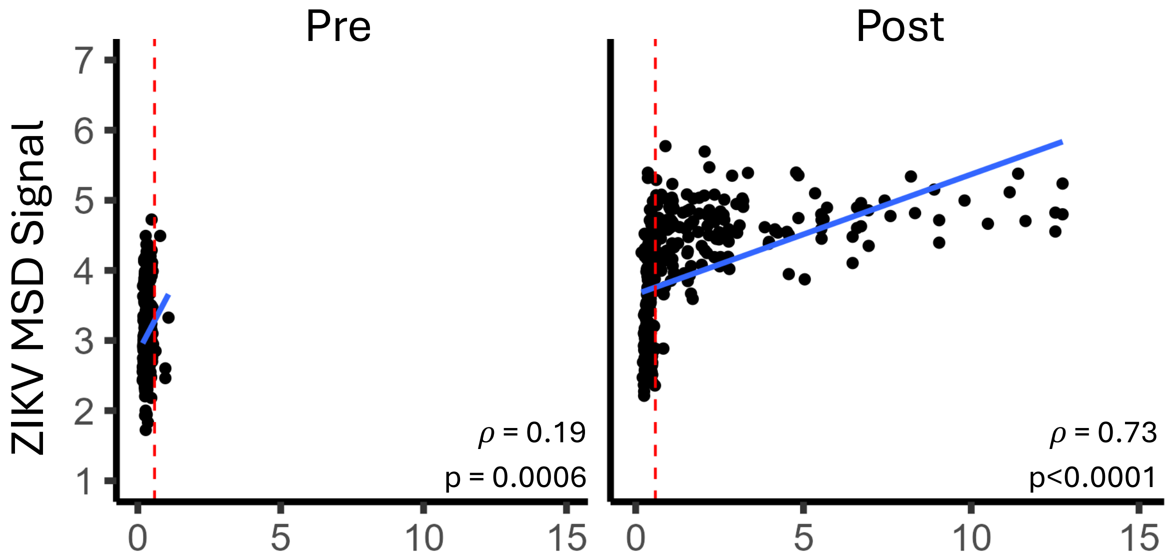
